# Dual exposure-by-polygenic score interactions highlight disparities across social groups in the proportion needed to benefit

**DOI:** 10.1101/2024.07.29.24311065

**Authors:** Sini Nagpal, Greg Gibson

## Abstract

The transferability of polygenic scores across population groups is a major concern with respect to the equitable clinical implementation of genomic medicine. Since genetic associations are identified relative to the population mean, inevitably differences in disease or trait prevalence among social strata influence the relationship between PGS and risk. Here we quantify the magnitude of PGS-by-Exposure (PGSxE) interactions for seven human diseases (coronary artery disease, type 2 diabetes, obesity thresholded to body mass index and to waist-to-hip ratio, inflammatory bowel disease, chronic kidney disease, and asthma) and pairs of 75 exposures in the White-British subset of the UK Biobank study (n=408,801). Across 24,198 PGSxE models, 746 (3.1%) were significant by two criteria, at least three-fold more than expected by chance under each criterion. Predictive accuracy is significantly improved in the high-risk exposures and by including interaction terms with effects as large as those documented for low transferability of PGS across ancestries. The predominant mechanism for PGS×E interactions is shown to be amplification of genetic effects in the presence of adverse exposures such as low polyunsaturated fatty acids, mediators of obesity, and social determinants of ill health. We introduce the notion of the proportion needed to benefit (PNB) which is the cumulative number needed to treat across the range of the PGS and show that typically this is halved in the 70^th^ to 80^th^ percentile. These findings emphasize how individuals experiencing adverse exposures stand to preferentially benefit from interventions that may reduce risk, and highlight the need for more comprehensive sampling across socioeconomic groups in the performance of genome-wide association studies.

## Introduction

As the discovery power of genome-wide association studies has increased to the point where meaningful stratification of risk for complex disease can be contemplated, attention is beginning to turn toward issues surrounding clinical implementation of polygenic scores (PGS)^1–5^. Two crucial concerns are the definition of appropriate thresholds, if any, guiding therapeutic intervention^6,7^; and ensuring that such thresholds are deployed equitably across population groups. The lack of portability across continental ancestry groups has already been the focus of considerable research^8,9^, but equally important is ensuring that healthcare is provided equitably across socioeconomic strata^10–12^. Since ancestry, ethnicity, and social determinants of health are inexorably linked, consideration of genetics in this context inevitably engages particularly vexing scientific and social issues.

Limited studies have provided empirical evidence of PGSxE interactions across environmental exposures in biobank scale datasets^13,14^, and consequently the overall contribution of PGSxE to the genetic architecture of disease has not yet been quantified. While additive effects of environmental exposures can be sufficient to markedly alter the relationship between PGS and disease prevalence or therapeutic outcomes, it is also important to discern whether genotype-by-environment interactions are prevalent enough to affect clinical implementation. Individual SNP-by-environment interactions are for the most part an order of magnitude smaller than main effects^15–17^, and consequently escape detection even in very large GWAS. However, thousands of such interactions can collectively lead to PGS-by-environment interactions that do meaningfully modify PGS accuracy^18,19^. Durvasula, et al.^14^ identified three categories of interaction, namely ones that increase the variance of effect sizes, ones that are correlated in direction of effect, and ones that induce PGS-exposure correlations^14^. Exposures may be cultural, behavioral, abiotic, biochemical, or other biological influences, such as sex which shows systematic amplification of SNP effect sizes for cardiometabolic traits in females as demonstrated by Harpak and colleagues^20^.

We have also drawn attention to a particular type of PGS-by-exposure interaction, (de)canalization, which results in enhanced or reduced magnitude of risk at the extremes of the polygenic score distribution^13^. Analysis of ten diseases and 150 exposures in the UK Biobank showed that this effect is pervasive and shows characteristic patterns that are for example quite different for obesity as measured by body mass index or waste-to-hip ratio. Under persistent stabilizing selection the genetic architecture may evolve to be buffered against genetic or environmental variation^21,22^, and the argument is that contemporary dietary and psychological exposures, for example, have disrupted this relationship^23,24^. Disease promoting exposures both increase the overall prevalence and the impact of high genetic risk, such that PGS have different predictive power across environments, including socioeconomic strata.

Here we systematically characterize the extent of PGS-by-exposure interactions for pairs of exposures in relation to seven disease classifications (type 2 diabetes and coronary disease, obesity (measured both as BMI and WHR), inflammatory bowel disease, chronic kidney disease, and asthma) in the UK Biobank^25^, the first four evaluated both as prevalent and 10-year incident cases. We identify specific key exposures exhibiting multiple interactions for each disease and show that there is a strong positive relationship between the excess disease variance explained due to interaction effects and amplification of SNP effect sizes. After using cross-validation to confirm the robustness of several interactions, we introduce the notion of the Proportion Needed to Benefit (PNB) as a metric for defining thresholds for clinical implementation. We conclude with discussion of whether calibration of PGS amounts to correction factors that could lead to biases that exacerbate or mitigate health disparities. Our analyses make a strong case that the transferability of polygenic scores across socioeconomic and other exposure strata is often poor^18^, and generally results in bias against accurate risk assessment in higher risk populations.

## Results

### Impact of polygenic scores on disease is highly context-specific

We begin by asking to what extent do combinations of environmental exposures modulate the disease risk vs PGS relationships? Leveraging the self-reported white-British subset of the UK Biobank data (n=408,801), we generated polygenic risk scores for seven common diseases, namely coronary artery disease (CAD), type 2 diabetes (T2D), body mass index (BMI)- and waist-to-hip ratio (WHR)-thresholded obesity, inflammatory bowel disease (IBD), chronic kidney disease (CKD) and asthma (see Methods). The disease risk was evaluated as both prevalent cases (denoting all self-reported cases and/or ICD10 codes) as well as incident cases (denoting incidence within 10 years of recruitment based on ICD10 codes and date of first in-patient diagnosis). Supplementary Table S1 contains the list of UK Biobank fields, ICD10 codes for each disease, GWAS summary statistics used for PGS and number of SNPs after pruning and thresholding. We then compared disease risk as a function of PGS with respect to pairwise-combinations of 75 environmental exposures (approximately ^75^C_2_ combinations) categorized into diet, lifestyle, socioeconomic factors, early life factors, metabolite levels and physiological factors including sex (Supplementary Table S2). Each exposure was split into two groups, namely, low vs high-risk based on the mean values for quantitative exposures and answers provided in the UKB questionnaire for categorical exposures. Thus, a combination comprising of two exposures has four levels of risk: low-risk in both exposures (E_00_, lowest overall risk denoted by yellow curve), high-risk in both exposures (E_11_, highest overall risk denoted by purple curve), and high/low-risk in one of the exposures (E_01_ or E_10_, blue or red curves).

Figure 1 depicts three scenarios showing the varied impact of PGS on the disease with respect to different combinations of environmental exposures. In each case, the x-axis is the percentile of PGS-CAD and the y-axis is the 10-year CAD incidence in the UK Biobank cohort. In the first case (Figure 1A), for the exposure pair ’fresh fruit intake along with number of days/weeks walked 10+ minutes’, no difference is observed with respect to four levels of environmental risk levels, i.e. low fruit intake and low physical activity, low fruit and high physical activity, high fruit and low physical activity and high fruit and high physical activity. In the second case (Figure 1B), we observed an additive impact of PGS with respect to clinical risk factors - triglyceride and cholesterol levels - as the curves depart, showing an incremental 2-3% increase in CAD incidence with exposure risk levels along the PGS spectrum, such that the two exposures have comparable effects on CAD risk. At the top percentile of PGS-CAD, comparison of individuals with high triglyceride levels and high cholesterol levels (purple curve, E_11_) implies an average offset of risk from 23% to 16% by reducing cholesterol levels (<6 mmol/L, orange curve E_10_) and further to 11% by additionally reducing their triglyceride levels (<1.7 mmol/L, yellow curve E_00_). We note that these curves establish an association and do not necessarily imply causality between exposures and disease risk, so the notion of offsetting risk by changing the exposure is hypothetical and should be considered illustrative only. The third case shows an increasingly large impact of sex and sex-adjusted testosterone levels on the CAD incidence where the deviations between high vs low-risk groups increases as the PGS increases (Figure 1C). Males with low testosterone levels (E_11_, purple curve) have the highest risk of CAD compared to females with high testosterone levels (E_00_, yellow curve) and the impact increases with the increase in PGS, such that the difference in incidence is 4.8% at the intermediate PGS and 19% in the top percentile of PGS.

**Figure 1:**
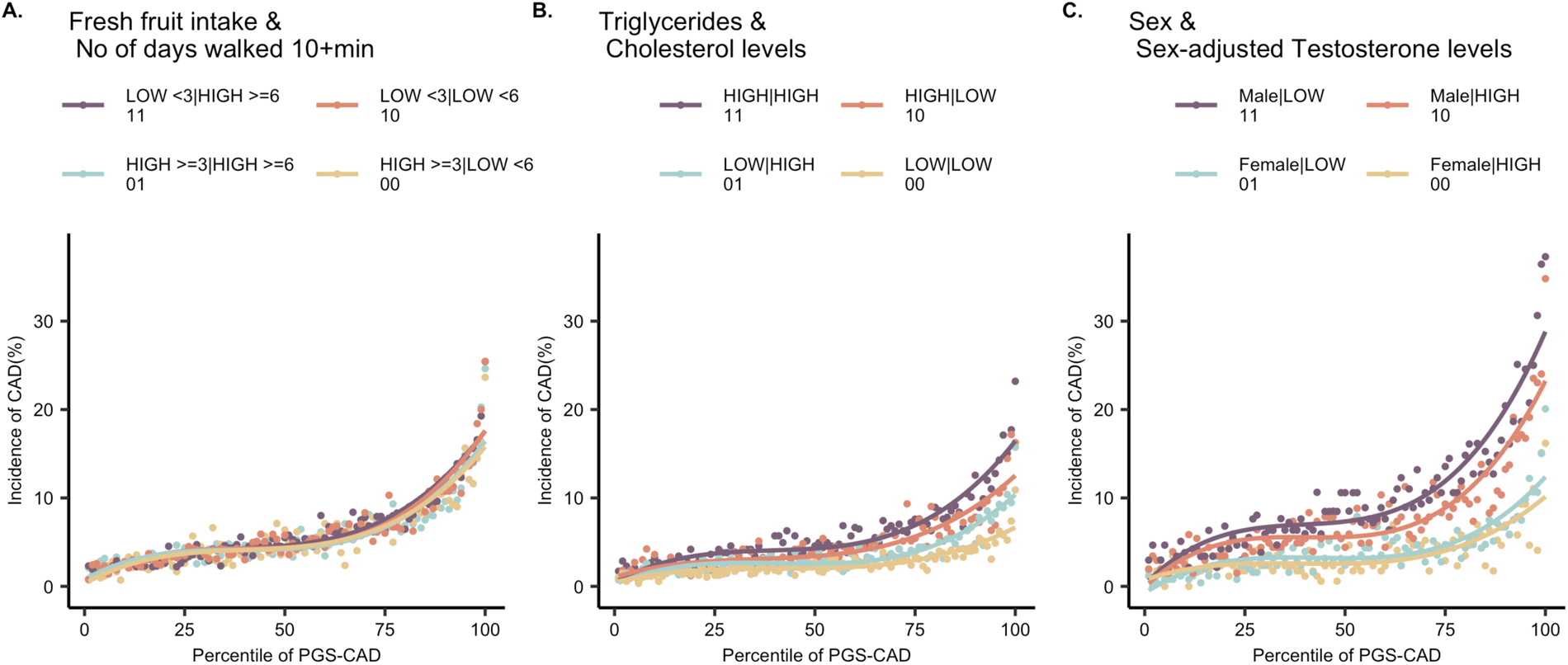
Varied impact of PGS-CAD on CAD incidence with respect to combinations of exposures. The curves show the relationship between incidence of CAD (%) vs percentile of PGS-CAD for combinations of exposures: **(A)** Fresh fruit intake and number of days walked 10+ minutes showing no effect with respect to high or low exposure risk levels, **(B)** Triglyceride and cholesterol levels showing a constant 2-3% shift with respect to different exposure risk levels, **(C)** Sex and sex-adjusted testosterone levels showing an increasingly large impact with respect to different exposure levels. Note that individuals taking cholesterol- or blood pressure-lowering medications were removed from all computations involving CAD risk.

Assessing the combinations of exposures has several advantages. Firstly, refinement of disease risk stratification with respect to pairs of exposures and genetic predisposition can be discerned. For example, in Figure 1C, males with low testosterone levels are uniformly at about 2-3% higher risk of CAD compared to males with high testosterone levels, irrespective of their PGS. Whereas, within females, high testosterone levels become protective only for women with high genetic predisposition to CAD (>75^th^ percentile of PGS). Assessing the PGS-risk relationships in the overall population without environmental stratification or even with single exposures averages out the effects of the contributing factors which may play a significant role in explaining the variability in disease risk observed among individuals. Secondly, it allows evaluation of the competing effects of the exposures in terms of their genetic influences on disease risk while controlling for the other environment. That is, we can ask whether two exposure effects are comparable, or one is more pronounced resulting in disproportionate increase in disease risk – and subsequently whether its influence might be offset by mitigating factors. For example, PGS has a larger impact on CAD with respect to sex (CAD risk at top percentile of PGS in males is 34.9%), compared to testosterone levels (risk at top percentile of PGS in low testosterone group is 29.0%). However, both factors modify the PGS-risk relationships such that high-risk in both the dimensions i.e. high genetic risk and high-risk exposure, becomes synergistically worse (risk at top percentile of PGS in males with low testosterone is the highest, 37.3%) (Supplementary Figure S1). Thirdly, quantifying how both factors contribute towards explaining the variance in the disease allows for gain in predictive accuracy of PGS in high-risk exposures. The prediction R^2^ of PGS in males with low testosterone levels is 15.4% compared to 11.2% in females with high testosterone levels and 12.5% in the overall population without any environmental stratification (Supplementary Figure S1). Despite the same PGS being evaluated against CAD risk for each of the three cases in Figure 1, the varying response across contexts underscores the complexity of interpreting these scores.

### Assessment of interaction between polygenic risk and exposures

We next asked whether the environmental effects are additive or whether certain combinations of exposures interact with PGS to exacerbate disease risk disproportionately thus modifying the PGS-risk relationships. We evaluated two models, (i) logistic regression with an interaction term on raw, non-binned data; (ii) liability threshold disease risk modeling to assess expected risk at each PGS percentile under the additive expectation. First, we tested for a significant interaction term of PGS with exposure-pair (p < 0.05) in the logistic regression model on the raw, non-binned data:

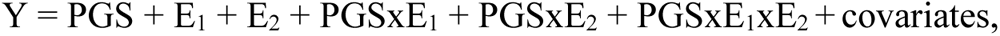

where Y is the case-control status of the disease (0/1), E_1_ is first exposure (0/1), E_2_ is the second exposure (0/1) and the covariates include age and five genetic PCs.

Next, we asked if the observed disease risk per percentile-PGS with respect to high vs low-risk exposures follows an additive expectation or PGSxE interaction. We utilized the liability threshold model to evaluate disease risk per percentile of PGS given the environmental effects are additive^13^ (see Methods). Assuming the underlying risk distribution in the overall population has a mean (μ) zero and variance (σ) 1, the liability threshold (*t*) can be determined from the prevalence in the overall population. Then, the mean of the underlying liability distribution (μ_i_) at each PGS percentile *i*, given the environmental effects are additive can be estimated:

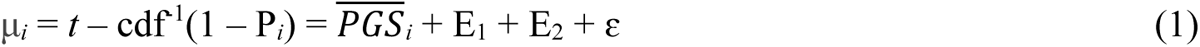

where, *t* is the liability threshold determined from the overall population prevalence, P*_i_* is the observed prevalence at PGS percentile *i*, 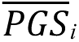 is the mean PGS at percentile *i*, E_1_ is the first exposure and E_2_ is the second exposure. Then, the expected prevalence (P*i*’) at each PGS percentile *i* is computed (assuming the variance of the underlying liability distribution (σ*_i_*) at each PGS percentile *i* is 1 under the null model) as:

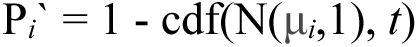

We computed the deviations of disease risk at the extremes of the PGS (> or < +/-2 σ) between high-risk vs low-risk exposures because we expect the deviations at the extremes will usually be associated with PGSxE influences on risk throughout the distribution. The metric to compute it is denoted as delta^13^ (see Methods). We identified significant interaction cases if the departure of delta_observed_ from delta_additive_ was greater than 2 standard deviation units. Across all 24,198 PGSхExposure models, 746 (3.1%) were significant by the above two criteria, at least three times more than expected by chance under each criterion (also Methods). Next, we evaluated the expected risk by including the interaction terms in the model i.e.

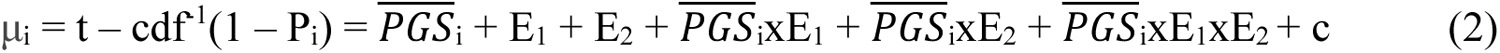

and computed the excess disease variance due to interaction effects as the ratio of the sum of signed effects from the interaction model i.e. β_PGS_ + β_PGSxE1_ + β_PGSxE2_ + β_PGSxE1xE2_ (equation 2) relative to β_PGS_ from the additive model (equation 1), following Darvusala and Price^14^.

Figure 2 shows three scenarios of interaction effects established using the above models (i) Low omega-6 fatty acid (ω6FA) levels and past tobacco smoking interact with PGS-CAD to exacerbate incident CAD risk (Figure 2A), (ii) High glucose and low ω6FA levels interact with PGS-T2D to exacerbate incident T2D risk (Figure 2B), (iii) High glucose and low ω3FA levels interact with PGS-BMI to exacerbate incident obesity risk (Figure 2C). The middle panel shows the expected risk (grey dashed curves) if the environmental effects were additive (equation 1). The excess disease risk in the observed data compared to additive expectation is captured by including the interaction between PGS and environments in the model (equation 2), shown by grey dashed-curves in the third panel. The adjusted R^2^ (reported in figure 2) of the interaction model shows significant improvement over the adjusted R^2^ of the additive model. This confirms that these exposures interact with PGS to increase disease risk non-additively. These models were also validated on independent test sets showing consistent findings i.e. improvement in R2 of interaction models relative to additive models (Supplementary Figure S2). Table 1 summarizes the departure of deviations at the extremes of PGS in the high vs low-risk exposure groups and excess disease variance due to interaction effects from the overall model for the above three cases.

**Figure 2:**
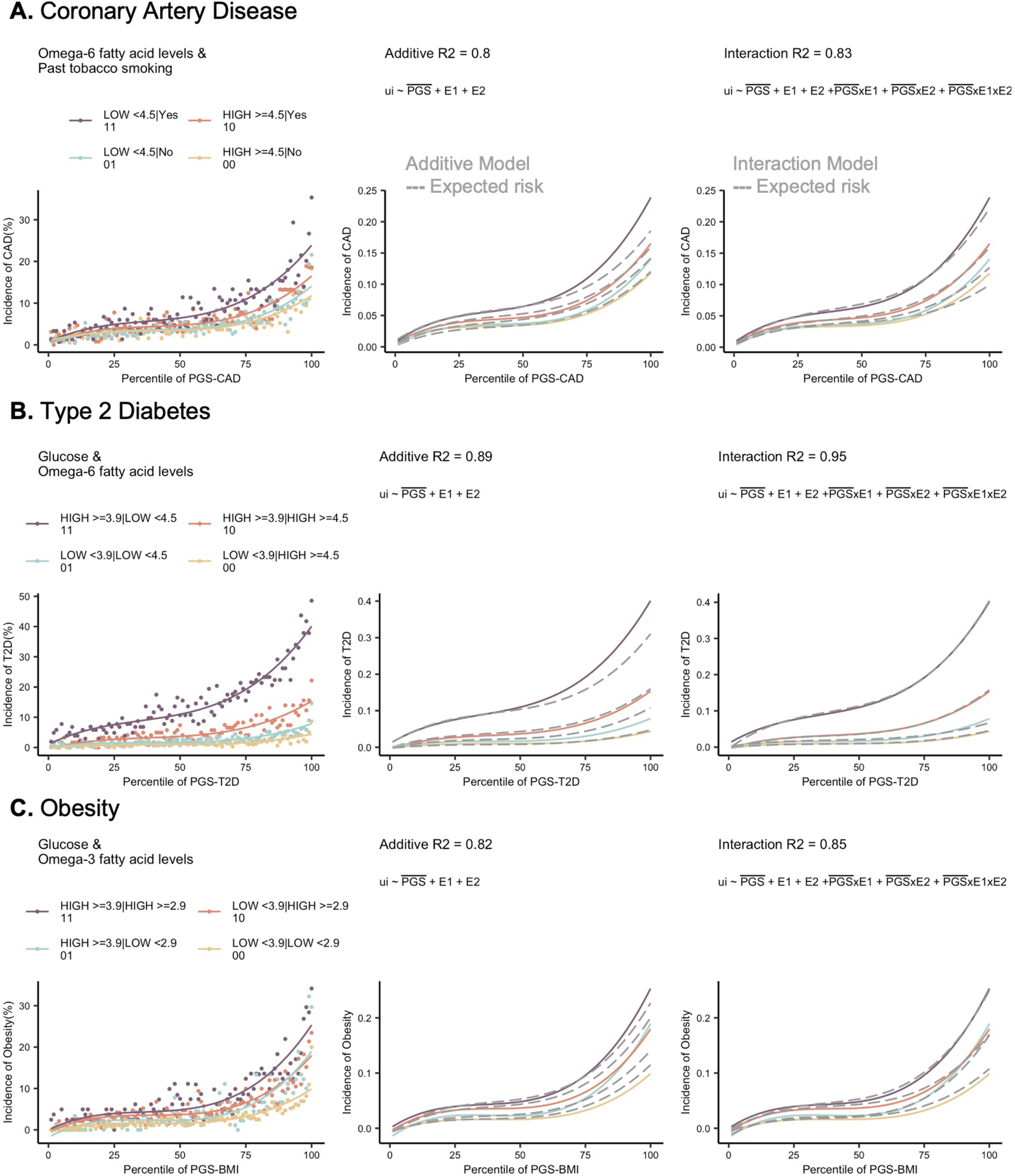
Assessment of interactions between PGS and exposures. **(A)** Incidence of CAD vs percentile PGS-CAD with respect to omega 6 fatty acids and past tobacco smoking, **(B)** Incidence of T2D vs percentile of PGS-T2D with respect to glucose and omega 6 fatty acid levels, **(C)** Incidence of obesity vs percentile of PGS-BMI with respect to glucose and omega 3 fatty acid levels. The second panel in each case shows the expected risk if the environmental effects are additive as shown by the grey dashed curves obtained using liability threshold disease risk modeling, overlaid on the observed curves. The third panel shows the expected risk (grey dashed curves) under the interaction model. The adjusted R^2^ of the additive and interaction models are reported on the top of each panel.

**Table 1:** PGSxE metrics for three cases: Coronary artery disease with respect to Omega-6 fatty acids and past smoking, type 2 diabetes with respect to glucose and omega-6 levels and obesity with respect to glucose and omega-3 fatty acid levels.

| Disease | Exposure | PGS<sub>extremes</sub> Model |  |  | Overall Model |
| --- | --- | --- | --- | --- | --- |
|  |  | Departure (sdu) | Delta observed (E<sub>11</sub>-E<sub>00</sub>) | Delta expected (E<sub>11</sub>-E<sub>00</sub>) | Excess disease variance due to interaction effects |
| CAD | Omega-6 and past smoking | 2.947 | 0.156 | 0.067 +/- 0.043 | 1.185 |
| T2D | Glucose and omega-6 | 2.188 | 0.332 | 0.274 +/- 0.058 | 1.287 |
| Obesity | Glucose and omega-3 | 4.644 | 0.237 | 0.083 +/- 0.049 | 1.221 |

Further, to ensure that the deviations in risk across PGS spectrum with respect to environmental exposures are not solely driven by prevalence/incidence differences or biased by PGS developed from GWAS of the overall population rather than exposure specific GWAS, we performed a series of sensitivity analyses described in the supplementary methods. These confirmed that there is a considerable tradeoff between less biased estimation of allelic effects in exposure-specific populations and increased estimation variance due to smaller sample sizes^26^. By equilibrating sample sizes and disease prevalence, and generating permutations of random dichotomous traits, we also show that prevalence differences associated with high genetic risk reflect synergistic interactions between exposures and polygenic risk independent of the influence of overall elevated prevalence in certain exposures (Supplementary Figures S3, S4 and Methods).

### Pervasive polygenic risk-by-exposure interactions (PRSxE) across common diseases

Across all disease-exposures, we find evidence of pervasive polygenic score-by-exposure interactions influencing common disease risk. Figure 3 shows the exposure pairs showing PGSxE for prevalent CAD (Figure 3A), incident CAD (Figure 3B), incident T2D (Figure 3C) and prevalent chronic kidney disease (Figure 3D) respectively and Supplementary Figure S5 shows the exposure pairs for the other diseases. These are established using the above-described models, namely comparison of (i) significant interaction term (p<0.05) of PGSxE regression on non-binned data, (ii) Departure of delta_observed_ from delta_additive_ greater than 2 standard deviation units, where delta is defined as the deviations of disease risk at the extremes of PGS (> or < +/-2 σ) between high vs low-risk exposure (see Methods). The width of the arc within each disease is proportional to delta_observed_ i.e. deviation in disease risk at the extremes of PGS between high-risk vs low-risk exposure, while the number of linkages per node denote multiple interactions exhibited by the key or major exposures for each disease.

**Figure 3:**
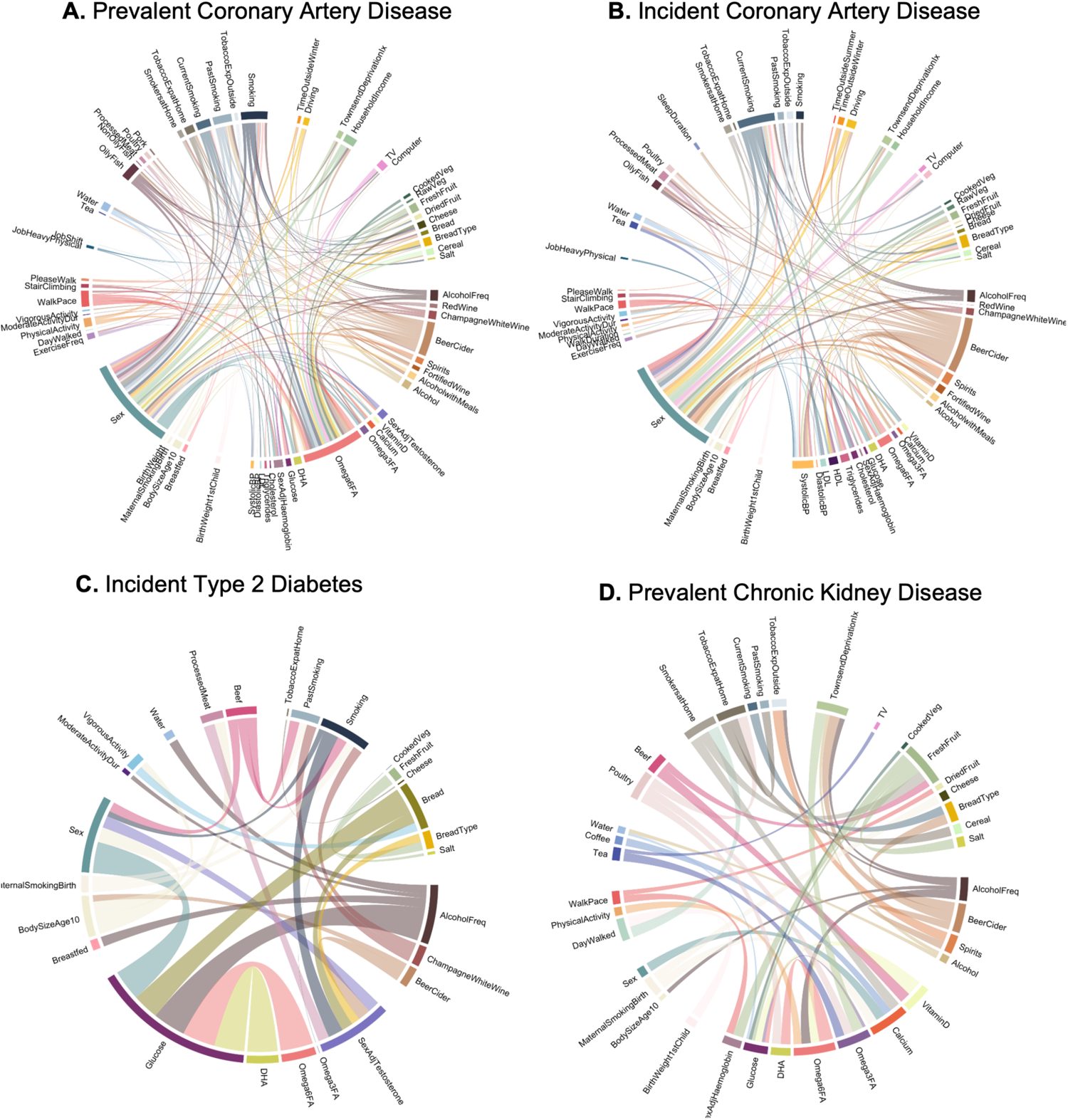
Combinations of exposures showing significant PGSxE for common diseases. **(A)** Prevalent coronary artery disease (CAD), **(B)** Incident CAD, **(C)** Incident type 2 diabetes (T2D), **(D)** Prevalent Chronic kidney disease (CKD). The width of the arc denotes the delta_observed_ i.e. deviations of disease risk at the extremes of PGS between high-vs low-risk exposure and the number of linkages per node denote the multiple interactions exhibited by key or major exposures for each disease.

For CAD, the incident cases (6% incidence) and prevalent cases (11.7% prevalence) show high concordance in the significant exposure pairs showing interaction effects (Figure 3A and 3B). The key exposures exhibiting multiple interactions include sex, weekly beer intake, ω6FA and smoking which in combination with other exposures interact with PGS to exacerbate CAD risk. For example, PGS has a significantly larger impact on CAD risk in males with slow walking pace, a measure of physical fitness (incidence at PGS_top_ = 30%) compared to females who walk briskly (incidence at PGS_top_ = 7%) (Supplementary Figure S6A). However, brisk walking in males offsets this risk, rendering it comparable to the overall risk observed in females with slower walking pace (incidence at PGS_median_ = 7%). Nevertheless, it is important to note that even with brisk walking, males exhibit elevated risk at higher PGS levels (> 75^th^ percentile). Reduced levels of polyunsaturated fatty acids (PUFAs) such as ω6FAs, ω3FAs and docosahexaenoic acids (DHA) have particularly adverse effects on CAD risk^27,28^, especially in people with high weekly beer and cider intake, and this is accentuated at elevated PGS-CAD levels (Supplementary Figure S6B). The risk curves for prevalent CAD cases show similar findings but even higher deviations of disease risk between high vs low risk exposures (Supplementary Figure S7 and Table S3).

Conversely, while clinical risk factors for CAD like cholesterol, LDL, HDL, triglycerides, and blood pressure are widely recognized, evidence demonstrates their additive effects when combined with polygenic risk scores (PGS)^29–31^. However, evaluating these risk factors within specific contexts unveils interaction effects with PGS, leading to elevated CAD risk which has clear implications for personalized medicine^32,33^. Thus, neither genetic risk nor other non-genetic risk factors such as clinical or environmental risk factors should be considered in isolation or modeled merely as additive contributions for disease risk evaluation. For example, high cholesterol has adverse effects on CAD risk particularly in smokers who have high genetic predisposition to CAD (Supplementary Figure S6C for incident CAD, S7C for prevalent CAD), as does low levels of HDL along with white bread intake compared to wholegrain or brown bread intake; and high systolic pressure in individuals whose jobs involve heavy physical work also exacerbates CAD risk along the PGS spectrum (Supplementary Figure S6D, S7D for prevalent CAD).

For incident T2D, the major exposures include glucose levels, ω6FA levels, sex-adjusted testosterone levels, sex, bread intake, alcohol frequency, smoking and body size at age 10 (Figure 3C). PGS-T2D imposes larger impact in individuals with reduced levels of PUFAs and high levels of glucose leading to elevated T2D risk as the PGS increases (Figure 2B). High levels of glucose also have adverse effects on T2D risk in individuals who consume more bread (incidence at PGS_top_ = 40%) compared to those who consume less bread (incidence at PGS_top_ = 30%); whereas there is no effect of bread intake in individuals with low glucose levels (Supplementary Figure S8A). Both high processed meat and beef intake have much more adverse effects on T2D in smokers compared to non-smokers at high PGS-T2D (Supplementary Figure S8B). With respect to testosterone levels, smokers with low testosterone levels are at a higher T2D risk compared to non-smokers with high testosterone levels (Supplementary Figure S8C)^34^. Early life factors such as having plumper body size at age 10 as well as high genetic predisposition to T2D imposes high T2D risk in males (incidence at PGS_top_ = 40%) compared to females whose body size is plumper at age 10 (incidence at PGS_top_= 10%) or thinner (incidence at PGS_top_ = 7%) (Supplementary Figure S8D). Most of these interactions are also observed for prevalent type 2 diabetes (Supplementary Figure S5A).

For incident and prevalent obesity defined by body mass index (Supplementary Figure S5D, F), the major exposures include walking pace, Townsend deprivation index, vitamin D levels and alcohol frequency, as well as the PUFAs ω6FA and DHA. By contrast, for prevalent obesity defined by waist-to-hip ratio(Supplementary Figure S5E), the major exposures include weekly beer consumption, systolic blood pressure, ω6FAs, sex adjusted testosterone levels, and pork/beef intake. Supplementary Figure S9 shows the examples of two combinations of exposures: (i) body size at age 10 and weekly beer intake, (ii) sex adjusted testosterone levels and bread intake which interact with PGS-BMI and PGS-WHR respectively to influence obesity risk. In both cases, being plumper at age 10 along with high beer intake and having low testosterone levels along with high bread intake, risk for obesity is high, however the responses to the PGSs differ. Notably, there is exacerbation of obesity risk as the PGS-BMI increases (Supplementary Figure S9A, B), however for PGS-WHR, the overall difference in top vs bottom percentiles of PGS is small, there is attenuation of obesity risk as the PGS increases. This is observed (i) in individuals who consume more beer and have plumper body size at an early age compared to lean body size (Supplementary Figure S9C); and (ii) in individuals who consume more bread, and the curves tend to converge for low vs high testosterone levels (Supplementary Figure S9D). This apparent canalization of disease risk for WHR and decanalization for BMI possibly reflects the contrasting roles of polymorphisms for BMI acting in the central nervous system and influencing human eating behavior vs those for WHR acting on metabolic traits considered to have been under stabilizing selection throughout primate if not mammalian evolution^13^.

Inflammatory bowel disease (IBD) is a chronic inflammatory disease of the gut arising due to dysregulated immune response to an environmental trigger in individuals with high polygenic load. The major exposures showing interaction effects include low hemoglobin levels, low cholesterol levels – LDL and triglycerides, white bread type, and systolic blood pressure. Supplementary Figure S5B and S10 shows notable examples of interactions of exposure pairs with PGS-IBD influencing IBD risk. Low LDL levels along with high triglycerides levels and high systolic blood pressure are associated with enhanced polygenic influences on the IBD risk (Supplementary Figure S10A, B). Evidence suggests that patients with IBD have impaired and altered lipid metabolism due to chronic inflammation and thus these factors may be the outcome of the disease rather than being causal^35^. Lower LDL levels may also be attributed to reduced diet and weight loss in IBD patients due to bowel symptoms. Studies have also demonstrated an elevated risk of cardiovascular outcomes in IBD patients despite low prevalence of cardiovascular risk factors such as cholesterol, termed as the CVD paradox^36–38^. Anemia is a particularly strong risk factor for individuals in the top tertile of polygenic risk, showing synergistic interaction with smoking status (Supplementary Figure S10C). Dietary factors such as preference for white bread over whole grain bread also exacerbates the polygenic influences on IBD risk (Supplementary Figure S10D), though we caution that cause and effect relationships are not clarified with this approach, and that for example reverse causation may be involved (IBD changes dietary preferences). There were insufficient incident cases to compare meaningfully with prevalent IBD.

For prevalent CKD, the major exposures include calcium levels, vitamin D levels, omega-3 and omega-6 fatty acids, hemoglobin levels, fresh fruit intake, weekly beer and exposure to tobacco smoke (Figure 3D and Supplementary Table S3). PGS-eGFR has an inverse relationship with CKD risk. Intriguingly, within females, high calcium levels tend to exacerbate CKD risk compared to low calcium levels at high genetic risk of poor eGFR, whereas within males, there is no difference in CKD risk with respect to high vs low calcium levels (Supplementary Figure S11A). High tea intake imposes higher risk for CKD at low PGS-eGFR, however elevated levels of ω3FAs do not offset that risk (Supplementary Figure S11B). On the other hand, reduced levels of ω6FAs along with high glucose levels tend to exacerbate CKD risk with about 3-4-fold increased risk at the low PGS-eGFR, compared to high levels of ω6FA and low glucose (Supplementary Figure S11C). Consideration of socioeconomic status as measured by Townsend deprivation index reveals a surprising result that higher alcohol intake appears to be protective against CKD^39^, with no discernible difference in CKD risk between high vs low socioeconomic deprivation. However, reduced alcohol intake has adverse effects with about 2% higher CKD risk in the high Townsend deprivation group along the PGS spectrum (Supplementary Figure S11D).

For Asthma, we found fewer interactions and more additive connections of the dietary or lifestyle factors with PGS. The major exposures include hemoglobin levels, type of working environment such as jobs involved heavy physical work or standing, and exposure to tobacco smoke. For example, PGS has a higher impact on asthma for individuals with high exposure to tobacco smoke at home and high triglyceride levels; on the other hand, females with low income show an overall higher risk than males with low income levels, however notably at high PGS-asthma, the high-income group in both males and females has an elevated asthma risk possibly pointing towards pollution levels or industrialization in high income or urban areas, or to ascertainment biases such as access to healthcare (Supplementary Figure S12A, B).

Supplementary Table 3 contains the metrics of the PGSxE models across significant exposures for each disease i.e., departure at PGS_extremes_, delta_observed_, delta_expected_ as well as the excess disease variance due to interaction effects from the overall model. Overall, we noted consensus between significant major exposures interacting with PGS identified for both prevalent vs incident disease cases. While the issue of PGS portability across ancestries has been a major focus in the literature, these results highlight the need to identify social determinants of health or contexts where PGSs and the interactions between these factors impose a larger impact on disease risk and thus could be more informative in terms of their clinical utility to ameliorate health disparities.

### Gain in prediction accuracy in high-risk environments

We next asked whether the predictive accuracy of PGS (R^2^, denoting the variance explained by a PGS for disease) varies in stratified high vs low-risk exposures as well as in composite exposure models that include the interaction between PGS and environments. We assessed the predictive accuracy of three models: (i) Low-risk exposures (E_00_): Disease ∼ PGS + covariates; (ii) High-risk exposure (E_11_): Disease ∼ PGS + covariates; (iii) Composite model with interaction: PGS + E_1_ + E_2_ + PGSxE_1_ + PGSxE_2_ and PGSxE_1_xE_2_ + covariates. The covariates included age and five genetic principal components.

Figure 4 shows the average R^2^ (measured on raw, un-binned data i.e. per individual rather than per percentile, hence lower than the R^2^ measures in earlier figures showing R2 from liability threshold disease risk modeling) across combinations of major exposures for each disease for the three models. In each panel, the dashed line indicates R^2^ for PGS in the overall population (without any environmental stratification or interaction effects). Overall, there is a gain in predictive accuracy in the poor or high-risk exposures compared to low-risk exposures and in many cases a further gain for the composite exposure model indicating that interaction effects between PGS and combination of exposures improves the variance explained for the disease risk. Notably, the PGS accuracy in the low-risk exposure is almost always lower than that observed in the overall population. Thus, disease promoting exposures both increase the overall prevalence and the impact of high genetic risk, such that PGS have different predictive power across environments, including socioeconomic strata. For incident CAD (Figure 4A and Supplementary Figure 13A for prevalent CAD), the R^2^ for males (in combination with other risk exposures) is 1.4-fold higher than that in females (average R^2^: males 0.148, females 0.103), and marginal improvement of 1.48-fold for the composite model with interaction term (average R^2^: 0.155). Glucose shows a larger impact for incident T2D, with 1.7-fold higher R^2^ in individuals with high glucose levels (R^2^=0.135) compared to those with low glucose levels (R^2^ = 0.081) and 2.2-fold for the composite model with interaction effects (R^2^=0.18) (Figure 4B and Supplementary Figure S13B for prevalent T2D). Similarly, slow walking pace almost doubles the predictive accuracy of PGS-BMI on incident obesity risk (R^2^ = 0.098) compared to brisk walk (R^2^ = 0.053) and the composite interaction model shows a further gain of 3.6-fold (R^2^ = 0.194) (Figure 4C), confirming the evidence of strong interaction effects established between physical activity and PGS-BMI^13,40–43^. For both alcohol intake frequency and Townsend deprivation index, the gain in predictive accuracy of PGS-BMI for obesity risk in high-risk exposures is ∼1.5 fold and greater than 2-fold in the composite model compared to low-risk exposures. Obesity defined by BMI threshold > 30 yields similar results (Supplementary Figure S13C), whereas for obesity defined by WHR > 0.9 (Supplementary Figure S13D), the average R^2^ values were much lower than BMI and similar in high vs low-risk exposures. However, with the interaction term, weekly beer, DHA and systolic blood pressure showed almost 3-fold gain while testosterone levels and lamb intake showed a 2-fold gain in R^2^. For prevalent CKD (Figure 4D), reduced levels ω6FA levels, hemoglobin levels, calcium and glucose show a gain in predictive accuracy, while there is marginal improvement in the composite interaction model indicating weaker interaction effects for CKD. For prevalent IBD (Supplementary Figure S13E), intriguingly, the gain in predictive accuracy is even greater than 2-fold in low levels of LDL, high levels of triglycerides and high systolic blood pressure compared to their corresponding low-risk exposures, although the absolute R^2^ values are lower compared to other diseases. However, we observed weak evidence of interaction effects for IBD, with reduced average R^2^ in the composite exposure model with interaction effects. Similarly, the average R^2^ values for asthma were much lower compared to other diseases (Supplementary Figure S13F).

**Figure 4:**
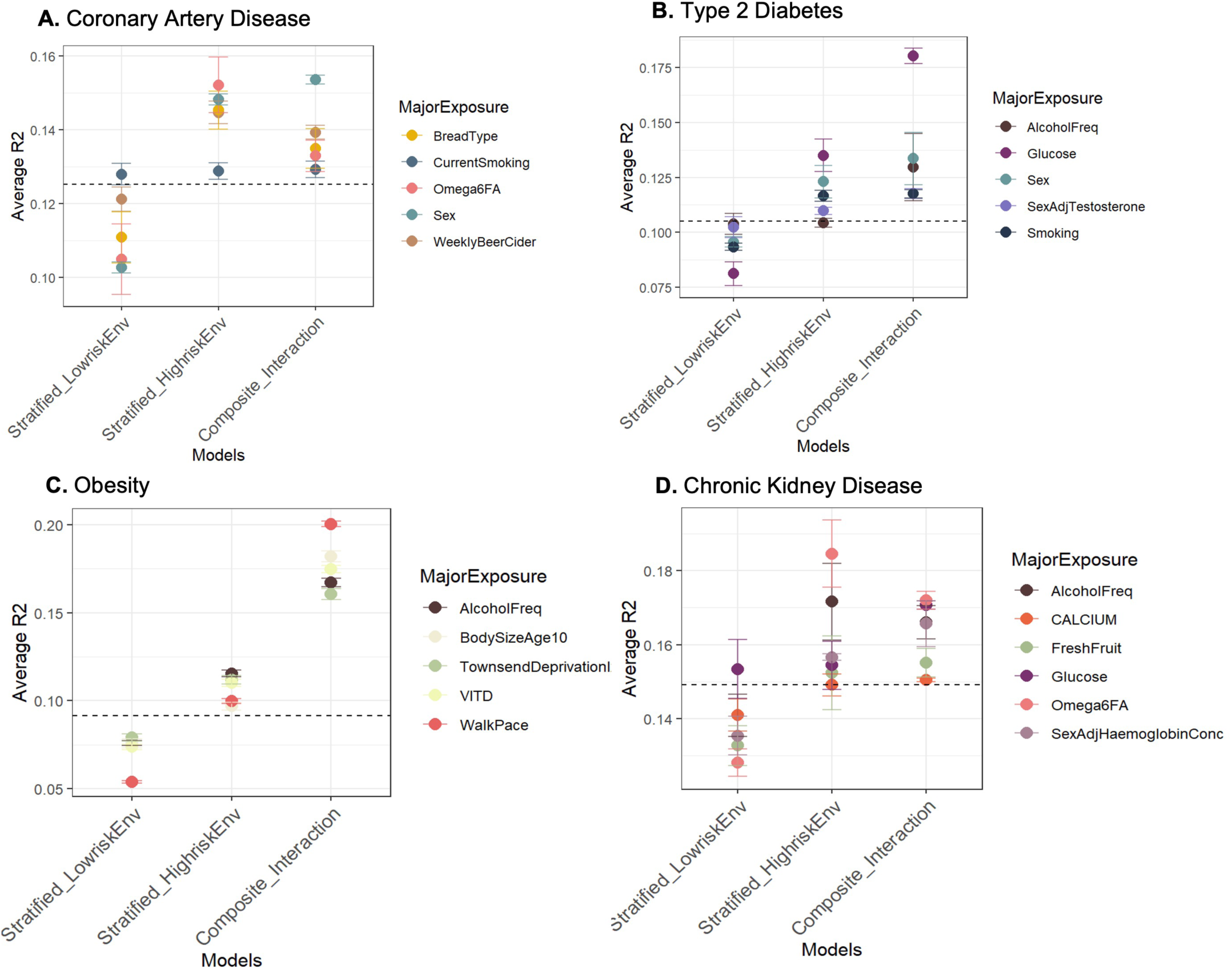
Gain in PGS prediction accuracy in high-risk exposures and composite exposure model including the interaction effects. The average R2 across significant exposure-pairs is shown for major exposures across each disease: **(A)** Incident coronary artery disease (CAD), **(B)** Incident type 2 diabetes (T2D), **(C)** Incident obesity with PGS-BMI, **(D)** Prevalent Chronic kidney disease (CKD).

The fold change in predictive accuracy observed here is comparable to that observed between between Europeans vs East Asians and Africans^8,44^, highlighting that the issue of portability must not only be considered with respect to ancestries but also across socioeconomic groups^18,45^ and environmental factors which modify genetic influences on health outcomes. It also highlights important factors for each disease that need to be considered while evaluating risk using PGSs. The difference in predictive accuracies is consistent with the difference in SNP-heritability (SNP-h^2^) across these exposures (Supplementary Figure S14) indicating there is an increase in genetic variance in disease promoting exposures, possibly leading to an excess number of disease cases or elevated risk in the poor environments as observed in PGS-risk relationships.

### PGSxE and its relationship with amplification

Given the pervasive evidence of PGSxE and gain in predictive accuracy of PGS observed in high-risk environments, we next sought to evaluate the mechanism of PGSxE by estimating the genetic effect sizes in the high vs low-risk exposures for major exposures identified for each disease. The exposure-specific genetic effects were computed for SNPs that were used for PGS construction (Supplementary Table S1), adjusting for age and genetic PCs. We next used multivariate adaptive shrinkage (MASH)^46^ to examine the mixture of covariance relationships (correlation and difference in magnitude) of SNP effect sizes along with their estimation noise^20^, between high-risk exposures (E_11_) vs low-risk exposures (E_00_). Across all exposure pairs, we observed that the exposure-specific genetic effects were almost perfectly correlated but systematically higher in magnitude in high-risk exposures with high predictive accuracies compared to low-risk exposures. This is consistent with the mechanism of amplification (rather than uncorrelated effects) explaining GxE i.e. systematic differences in numerous genetic effects, recognized by previous studies^14,18,20,47^. The amplification of genetic effects parallels the increase in variance of genetic effect sizes in the high-risk exposures compared to low-risk exposures consistent with the elevation of SNP-h^2^ supporting PGSxE effects. For example, for CAD, the majority of the genetic effects are nearly perfectly correlated but up to 2x higher in magnitude for smokers with reduced levels of ω6FAs compared to non-smokers with higher levels of ω6FAs (Supplementary Figure S14A). Further the SNP-h2 is also 8% higher in smokers with reduced levels of ω6FA compared to low-risk exposure (Supplementary Figure S14B). Thus, the genetic effects may be highly correlated across exposures but amplified in the high-risk environment leading to an increased genetic variance (SNP-h^2^).

Across the major exposure-disease combinations showing PGSxE, we find evidence of extensive amplification of genetic effects in the high-risk exposures. We computed the net amplification effect^20^ as the percentage of genetic effects with magnitude higher in high-risk exposure (E_11_) compared to low-risk exposure (E_00_) i.e. H > L minus percentage of genetic effects with magnitude higher in the low-risk exposure compared to high-risk exposure i.e. H<L for each exposure-pair. Figure 5 shows a strong positive correlation between the net amplification effect with the excess disease variance due to interaction effects from PGSxE models for prevalent CAD (r=0.63, p=4.8e-12, Figure 5A) and incident CAD (r=0.61, p=5.4e-10, Figure 5B). The significant positive correlation is consistent across all diseases (Supplementary Figure S15): prevalent T2D (r=0.47, p=0.002), incident T2D (r=0.67, p=.002), incident obesity (r=0.32, p=0.005), obesity-BMI (r=0.48, p=0.002), obesity-WHR (r=0.7, p=7e-06), prevalent IBD (r=0.59, p=0.045).

**Figure 5:**
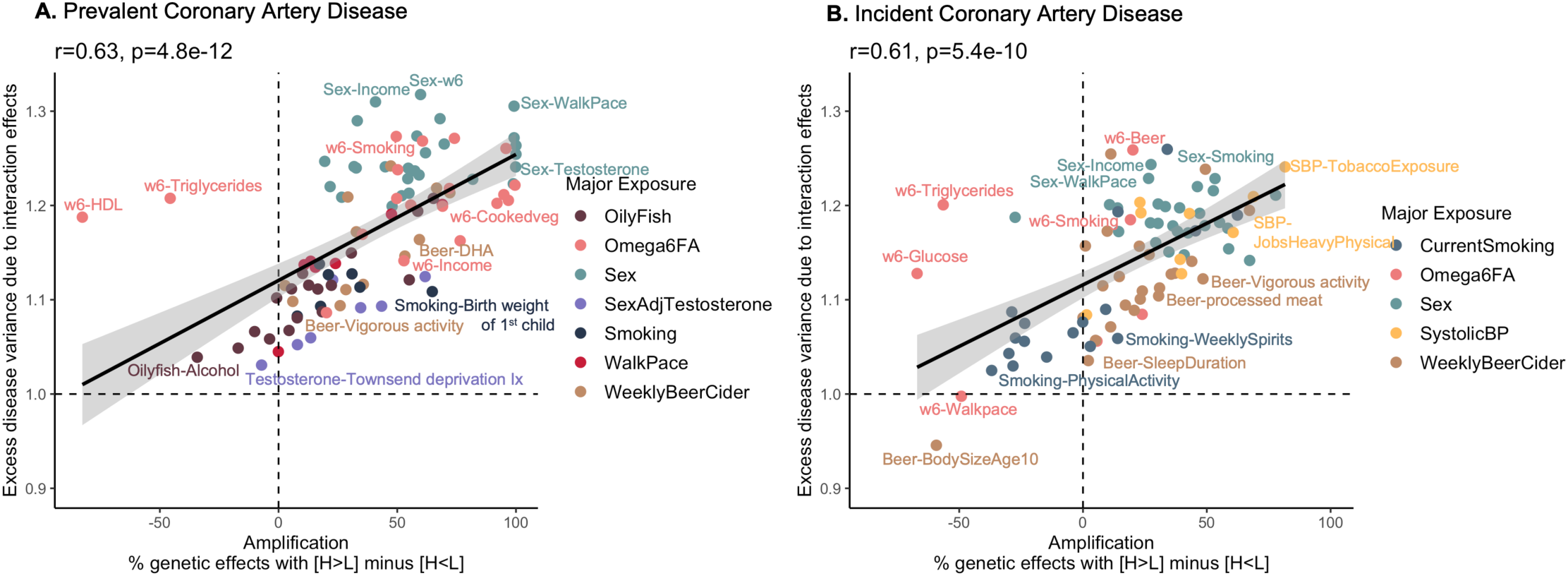
PGSxE shows a positive relationship with amplification of genetic effects. The x-axis denotes the net amplification effect (%) computed as the percentage of genetic effects with H (E_11_) > L (E_00_) minus percentage of genetic effects with H (E_11_) < L (E_00_). The y-axis denotes the excess disease variance due to interaction effects computed from the liability threshold disease risk modeling.

For CAD (Figure 5), sex interactions show the highest net amplification where genetic influences on CAD tend to be more pronounced in males than females along with other contributing factors such as low income, slow walk pace, low DHA and testosterone levels. This is true of both prevalent and incident disease, and beer consumption is also generically amplifying. In contrast, the genetic influences of reduced levels of ω6FAs are typically more pronounced in prevalent than incident CAD, with the exception of reduced ω6FA with high triglycerides where there is apparent suppression of genetic effects despite the high variance explained due to interaction effects. Interestingly, systolic blood pressure only appears to amplify genetic influences on incident disease. For type 2 diabetes, high glucose levels have amplified genetic effects along with contributing factors such as reduced ω6FA levels, high cholesterol, alcohol, and bread intake, but the magnitude of the amplification is typically greater for incident disease whereas the variance due to interaction effects (degree of decanalization^13^) is greater for prevalent disease (Supplementary Figure S15A,B). Omega 6 fatty acids only promote amplification for prevalent diabetes, implying that their influence is across the lifespan. Incident obesity is notable for relative suppression of genetic effects for adults who self-reported as being plump as children, which exemplifies a trend for canalization to associate with reduced effect sizes overall. Interactions with walk-pace contribute to the incidence of obesity apparently with little impact on effect sizes of variants contributing to the PGS (Supplementary Figure S15E). Low Townsend deprivation index, low testosterone levels and low vitamin D levels also show high amplification but moderate interaction effects. Obesity defined by BMI and by WHR show very different patterns of amplification Supplementary Figure S15D, F), although in both cases the significant positive relationship between amplification and variance due to interaction effects is maintained. Supplementary Table S4 contains the net amplification effect (%) for significant exposures across diseases.

### Consideration of PGSxE to assess the Proportion Needed to Benefit (PNB)

One of the major barriers preventing the adoption of PGSs is challenges surrounding its clinical implementation. PGSs offer a significant value in risk stratification rather than overall prediction or diagnosis. Thus, PGS-based approaches may offer clinical utility in screening strategies, which when integrated with environmental and clinical risk factors enhance the ability to identify individuals on whom preventative interventions would be most impactful^4,48^. With pervasive evidence of PGSxE interactions influencing disease risk, risk thresholds can be identified to maximize the proportion of population that might benefit from lifestyle changes, while optimizing the risk reduction as well as response to interventions^6,7^. Here we introduce the notion of the Proportion Needed to Benefit (PNB) as a metric for defining thresholds for clinical implementation. Building on the number needed to treat (NNT)^6,7,49^ concept, the proportion needed to benefit is a measure of how many people need to reduce their high-risk exposure (modifiable diet or lifestyle in many cases) in order to benefit one person at increments along the polygenic score percentile range. It is computed as the inverse of the difference between area under the risk-PGS curve for high-risk exposure (E_11_) minus low-risk exposure (E_00_) at different PGS thresholds, scaled by the proportion of population sampled at each threshold.

**Figure 6:**
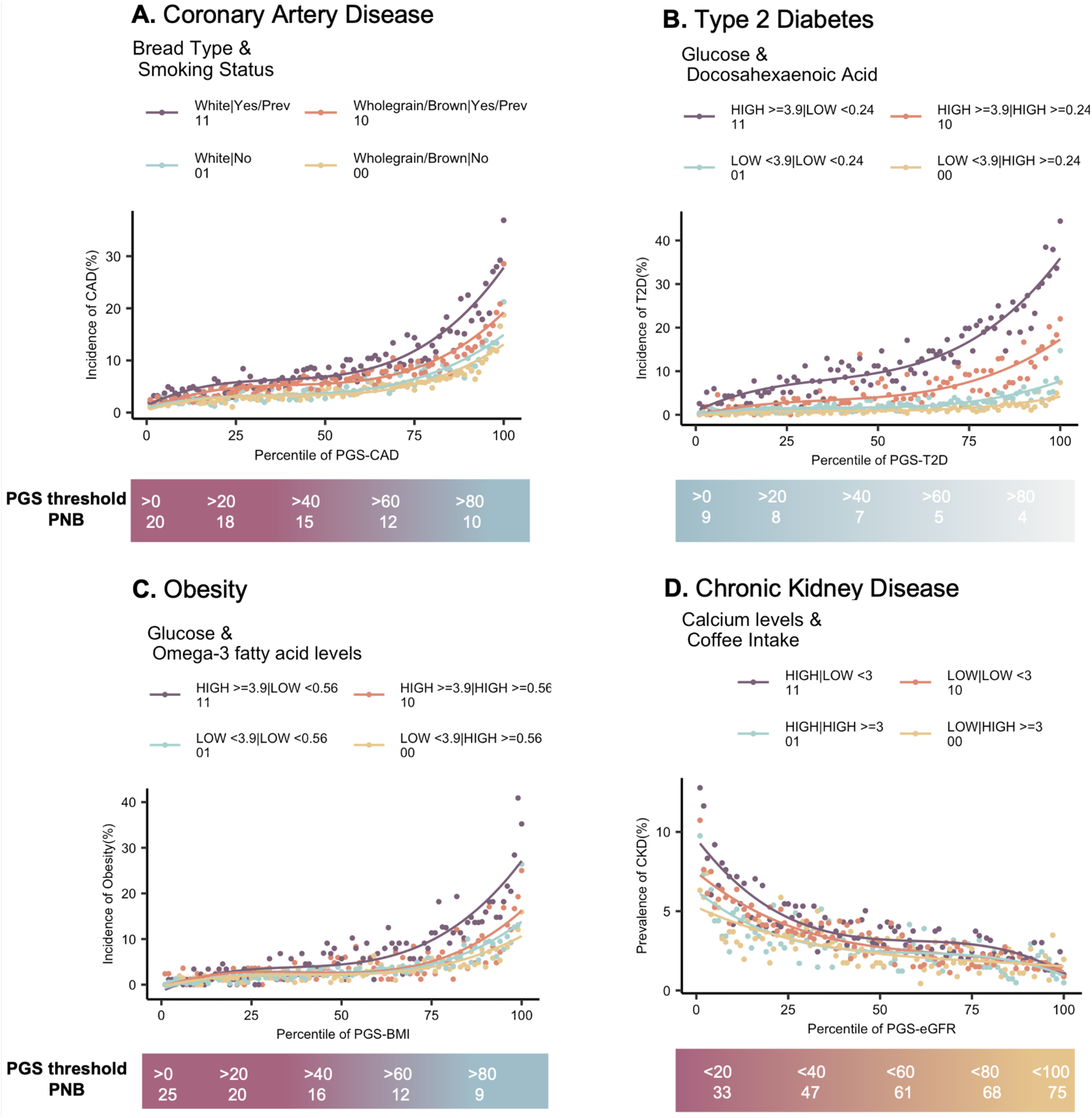
Proportion needed to benefit as a function of PGS thresholds in the high-risk modifiable exposure. In each case, the scale below the risk-PGS curves denotes the proportion needed to benefit (PNB) from high (E_11_) to low-risk (E_00_) exposure at increasing PGS thresholds for **(A)** Incident coronary artery disease (CAD), **(B)** Incident type 2 diabetes (T2D), **(C)** Incident obesity **(D)** Prevalent chronic kidney disease (CKD).

Figure 6 shows four examples of risk-PGS curves with respect to modifiable exposures along the scale denoting the PNB at PGS thresholds from left to right, PGS > 0 (overall population, without consideration of genetics), PGS percentile >10, PGS percentile >20 and so on. For example, with increasing PGS thresholds, the incidence of CAD increases in both high-risk exposure (E_11_) i.e. smokers who consume white bread as well as low-risk exposure (E_00_) i.e. non-smokers who consume wholegrain or brown bread (Figure 6A). However, targeting the entire population for lifestyle intervention without the consideration of genetic risk (i.e. PGS percentile >0) yields proportion needed to benefit as 20 implying 1 in 20 i.e. 5% of 40368 individuals (3863 CAD cases) who are in the high-risk exposure would benefit by reducing their CAD risk. Whereas, targeting the top 20% of the population based on their genetic risk (PGS percentile > 80) doubles the gain in benefit, yielding a PNB of 9 implying that 1 in 9 i.e. 11% of a smaller proportion of population (n=4037, CAD cases = 108) may benefit. On the face of it (see Discussion below), individuals in the high genetic risk and high-risk exposure (smokers consuming white bread) may reduce their CAD risk from 20% to as low as non-smokers who consume wholegrain bread (CAD incidence 9%) by switching their lifestyle. Consequently, the precision or positive predictive value increases with increasing PGS thresholds in both high and low-risk exposure but is 2-fold higher in the high-risk exposure (Supplementary Table S5), highlighting the effectiveness of targeting a smaller proportion of population stratified by both PGS and environmental risk to maximize the benefit from interventions. Similarly, Natarajan et al^50^ identified a high genetic risk group who benefitted from statin therapy with an NNT of 28, compared with 77 for the entire trial participants. Importantly as well, the negative predictive values are higher in the low-risk exposure and decreases with increasing PGS thresholds (Supplementary table S5) suggesting that unnecessary interventions could be avoided in the lower genetic risk groups^6^.

Similarly, 25% (1 in 4) individuals with high PGS-T2D (>75^th^ percentile) can in theory reduce their T2D risk by switching to a diet with lower blood glucose and elevated Docosahexaenoic acid (DHA) levels (Figure 6B). For obesity (Figure 6C), reducing glucose and increasing ω3FAs through diet may reduce obesity risk for 1 in 25 individuals (4%) in the entire population but at high genetic risk, PGS > 80^th^ percentile, 1 in 9 individuals (11%) may benefit. For CKD (Figure 6D), there is an inverse relationship between PGS-eGFR. Thus, in the overall population, PGS < 100^th^ percentile, 1 in 75 (13%) individuals may benefit by reducing calcium levels and increasing coffee intake, whereas at PGS < 20^th^ percentile, 1 in 33 (30%) individuals may benefit. Across all significant exposures vs diseases, the relative gain in benefit of 50% was attained at PGS thresholds in the 70^th^-80^th^ percentile. Overall, these results highlight the need to advance towards exposure-informed clinical implementation of PGSs and identify social strata or environments where PGSs impose greater impact on the disease.

## Discussion

Our investigations highlight the pervasive context-dependency of polygenic scores. The findings have implications from understanding the malleability of genetic architectures across environments, to implementation of PGS in personalized medicine, particularly in relation to promotion of equity in healthcare. Much has been written, appropriately, about the non-transferability of PGS across ancestry groups^10^, due mostly to drift in allele frequencies and linkage disequilibrium. However, the magnitude of PGS-by-exposure interactions places this discussion in the broader context of transferability of scores across populations with very different social determinants of health^18^.

To illustrate this, consider the case of chronic kidney disease (CKD). This high-mortality condition has attracted consequential debate over the use of so-called race correction factors to adjust for differences in prevalence between Blacks and non-Blacks^51^. Most initial diagnosis is by way of screening for serum creatinine, high levels of which are a biomarker for low estimated glomerular filtration rate and hence impaired kidney function. Since adult African Americans on average have twenty percent higher serum creatinine, for reasons that may or may not relate to kidney function, they are correspondingly more likely to be diagnosed with CKD^52^. Proponents of adjustment for this difference argue that statistical corrections are commonplace in medical assessment, and that it reduces expensive over-diagnosis in Black populations while promoting access to potentially limited dialysis for non-Blacks^53^. Opponents have successfully argued for cessation of the practice in the US on the basis that it is racist and stigmatizing and potentially leads to under-diagnosis. Millions of lives are affected by policy, and while our analyses do not bare directly on the question (since they were restricted to the White British subset of the UK Biobank cohort to avoid such complications), they place it in the broader context of health equity in at least four ways.

i. There are dozens of combinations of exposures that identify subsets of a quarter of the population whose risk for CKD is more than double that of the remainder, a difference that dwarfs that attributed to race. Some of the exposures are lifestyle-related and some are biochemical, and most are correlated with ancestry and ethnicity. This observation alone raises questions about screening and interpretation of kidney disease across socioeconomic, and other, groups.
ii. The accuracy of PGS is meaningfully greater in the high-risk exposure groups, in so far as much more of the variance in disease is explained by the PGS in high exposure environments. This finding is consistent across all diseases and relates to the amplification of genetic effects in individuals with environmental exposures that promote disease. Yet the vast majority of GWAS are performed in relatively affluent urban settings. While the signals seem to replicate well across populations, their cumulative effects on the PGS may not, which argues strongly for more inclusivity in genetic studies across the full range of social strata.
iii. The effect of exposures on the prevalence-PGS risk curve varies widely, often exhibiting synergistic amplification of the risk for individuals with high genetic and exposure risk. We have previously described this phenomenon as decanalization^13^. For CKD it is for example notable that prevalence only increases in for individuals in the top quartile of genetic risk for people with high tea intake (Supplementary Figure S11B). BMI shows more decanalization than WHR and interacts with somewhat different exposures, likely reflecting differences in long-term stabilizing selection on cognitive and metabolic aspects of weight gain^13^. In any case, the PGS is only one part of the equation for risk evaluation. Adjustment of population differences in PGS for example by centering the score^1,2^ do little to adjust for environmental impacts and may be as inappropriate as race correction factors.
iv. The consequences for personalized genomic medicine cannot be understated. While much focus is on positive prediction, we also note the implications for negative predictive values which are statistically higher in the low genetic risk subsets of low exposure-risk populations, often exceeding numbers needed to treat over 100. On the other hand, reduced negative prediction for those experiencing poor social determinants of health represents another potential issue for health equity.

Some of these conclusions are affected by a major limitation of this study, which is that we are for the most part unable to assess the causality of specific exposures. There are many possible explanations for genotype-by-environment interactions, some mechanistic and some statistical biases, including the common instance of genotype-environment correlations^54^: it is quite possible for PGS to be on average higher or lower in different socioeconomic groups because of historical legacies, for example. The high correspondence between prevalence and incidence relationships with environment is consistent with causality, but those estimates are confounded (a third or more of prevalent cases are also incident) and there may be other explanations. For example, some measures that we call exposures, particularly those related to exercise and diet, may represent rapid behavioral responses to the onset of disease. Furthermore, even if an exposure is causal, it may not be modifiable, if the damage due to the exposure is not reversible. While we identify dozens of possible lifestyle interventions that on the face of it would halve the prevalence of conditions as diverse as CKD, diabetes, and inflammatory bowel disease, it is not clear that they would be effective even if people heeded the epidemiological evidence. Methods such as Mendelian randomization may shed light on causality, but the complex covariance of so many of the exposures considered here urges caution in interpreting such studies.

Nevertheless, we would argue that a key implication of our findings is that exposures need to be considered when implementing PGS in settings where the intervention is expensive, has limited availability, or may be associated with detrimental side-effects. While PGS are sold as a key to personalized medicine, the fact is that the major benefits accrue at the population level: interventions with NNT between 5 and 10 benefit a small fraction of individuals but reduce incidence overall with great consequence. Considering cardiometabolic diseases, there is little argument for stratifying administration of drugs like statins and metformin by genetic risk, but there would likely be benefit to doing so for new generation drugs such as GLP-1 agonists^55,56^. For those, the proportion needed to benefit set by cutoffs of the PGS is critically a function of the exposure, with individuals who have high genetic risk differing in their likelihood or response by as much as five-fold or more. Assessment of the utility of exposure-informed clinical implementation of PGS to target expensive and limited therapies to those most likely to benefit need to be evaluated in a clinical trial framework.

Finally, a further limitation of our research is that most of the interactions have not been validated. We are replicating key results in the All-of-Us^57^ study but note first that the UK and US cohorts are demographically very different, and second that the self-reported exposure measurements in the two studies are not the same. Exact replication is not to be expected given these considerations as well as the pleiotropic nature of exposures that generally also have a high (but variable) correlation structure. Yet the overall pattern of context-dependence certainly is a general phenomenon, and it is in our view important that this perspective be prominent when the potential of polygenic risk evaluation is communicated to patients and to the general public.

## Methods

### Study population

The UK Biobank (UKB) is a large population-based cohort consisting of ∼500,000 individuals, recruited between 2006 and 2010 at 22 assessment centers spread across the UK^25^. The participants aged 40–69 at recruitment, completed baseline questionnaires about lifestyle, physical measures, medical history, and general health, as well as providing biological samples (blood, urine, and saliva) for genetic, proteomic, metabonomic analyses, and biomarker identification. In addition, the UKB has also generated data fields to indicate the first occurrence of a set of diagnostic codes for a wide range of health outcomes across self-report, primary care, hospital inpatient data, and death data, all mapped to a three-digit code of International Classification of Disease (ICD-10). In this study, we downloaded the genotype and phenotype from the UKB under application number 17984. The imputed genotype data (named v3) released in May 2017 for ∼ 96 million markers was downloaded. After selecting bi-allelic variants with imputation score > 0.9, minor allele frequency >1%, Hardy–Weinberg equilibrium *P* > 10^−10^ and <5% missing rate, a total of 8,063,507 SNPs was available for analysis. For the inclusion criteria of the participants, we selected a total of 408,801 self-reported White British individuals, (QC parameter ‘in.white.British.ancestry.subset’ == 1 as provided by the UKB), which were of similar genetic ancestry based on the principal component analysis of the genotype data and passed the sample QC filters^25^. Further, we utilized the first five principal components provided in the genotype QC data of UKB to account for population structure within this set of individuals in our statistical models. We excluded individuals who had withdrawn from the UKB by the time of the analyses and only included individuals for whom trait and environmental exposure values were reported.

### Phenotype data

The phenotype data was extracted for seven diseases and traits – coronary artery disease (CAD), type 2 diabetes, chronic kidney disease, obesity (relative to body mass index (BMI) and waist-to-hip ratio (WHR)), inflammatory bowel disease (IBD) and asthma. The prevalent cases included both self-reported and ICD10 codes. Whereas the incident cases included only ICD10 codes where date of first in-patient diagnosis-ICD10 was within the 10 years of recruitment in UKB i.e. year of first in-patient diagnosis minus year of recruitment >=1 and <= 10 years. For thresholded obesity, BMI >= 30 and WHR >= 0.9 was used. Supplementary Table S1 contains the complete list of UKB field IDs, ICD10 codes used, exclusion criteria, prevalence and incidence for each disease and trait.

### Environmental exposures

We utilized 75 environmental exposures categorized into diet, lifestyle, socioeconomic factors, early life factors, metabolite levels and physiological factors including sex (Supplementary Table S2). For each exposure, baseline measurements were taken. Each exposure was then split into two groups, namely high vs low based on the mean values of quantitative exposures and answers provided in the UKB questionnaire for categorical exposures. Individuals with missing phenotypic values, or who answered “Do not know” or “Prefer not to answer” were excluded. For sex, individuals whose genetically assigned sex (field ID: 22001) matched to the self-reported sex (field ID: 31) were included. For blood biochemistry biomarkers – cholesterol (field ID: 30690), triglycerides (field ID: 30870), LDL (field ID: 30780), HDL (field ID: 30760), diastolic blood pressure (field ID: 4079) and systolic blood pressure (field ID: 4080), we excluded individuals taking cholesterol-lowering or blood pressure medications (field IDs: 6153, 6177). Both testosterone levels (field ID: 30850) and hemoglobin levels (field ID: 30020) were adjusted by sex before generating high vs low groups for these exposures.

To assess PGSxE across combinations of exposures, we generated pairwise combinations of these exposures (approximately n=^75^C_2_ combinations). Thus, a combination comprising of two exposures has four levels of risk, namely low-risk in both exposures (E_00_, lowest overall prevalence denoted by yellow curve), high-risk in both exposures (E_11_, highest overall prevalence denoted by purple curve), and high/low-risk in one of the exposures (E_01_ or E_10_) (Supplementary Table S2). For consistency, high-risk in all cases was defined simply as the environment with the elevated risk overall and is not taken to imply that one or the other exposure is intrinsically high-risk. In some cases, notably alcohol consumption, this definition results in the same exposure being high- or low-risk for different conditions. Exposures with small sample size where four levels of risk were not attained were excluded.

### Computation of polygenic scores

Supplementary Table S1. provides the references of the independent European-based GWAS summary statistics used for each disease and trait and the number of SNPs after pruning used for the computation of polygenic score (PGS) on the UKB cohort. First, we subset SNPs with p < 0.001 from each GWAS summary statistic file and then performed LD pruning using ‘--indep-pairwise’ function in *PLINK*^58^ to filter out variants with LD R^2^ > 0.2 within a 1000 kb window. The ‘–score’ function in *PLINK* was used to obtain the weighted sum of the genotypes by log of odds ratios for diseases and effect sizes for traits to generate PGSs on the UKB cohort. We further also validated our results using a genome-wide PGS constructed using a Bayesian-based approach of estimating posterior SNP effect sizes under continuous shrinkage, PRS-CS^59^.

### Assessment of polygenic score by environmental interactions (PGSxE)

To assess PGSxE interactions, we evaluated two models – (i) logistic regression with an interaction term on raw, non-binned data; (ii) liability threshold disease risk modeling to assess expected risk at each PGS percentile under additive expectation. First, we tested for a significant interaction term of PGS with exposure-pair (p < 0.05) in the logistic regression model on the raw, non-binned data:

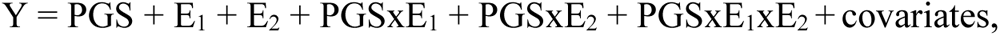

where Y is the case-control status of the disease (0/1), E_1_ is first exposure (0/1), E_2_ is the second exposure (0/1) and the covariates include age and five genetic PCs. Of the total 24,198 models assessed across all diseases, 5469 (22.6%) exposure-pairs showed significant interaction term (PRS xE_1_xE_2_) at p-value < 0.05, i.e. up to 4.5-fold more than expected by chance.

Next, we asked if the observed disease risk per percentile-PGS with respect to exposures follows an additive expectation or PGSxE interaction. We compared the shapes of risk-PGS curves, given that the underlying PGS have similar distributions across environmental risk levels. The prevalence/incidence of disease vs percentile of PGS curves with respect to combinations of environmental exposures were generated, each combination consisting of four levels by mean prevalence in each, namely, E_00_ (low-risk, lowest prevalence), E_01_, E_10_, E_11_ (high-risk, highest prevalence). Exposure-pairs with smaller sample size leading to zero prevalence in the lowest ten PGS percentile bins per group were removed. We utilized our recently developed liability threshold modeling approach^13^ to model expected disease risk (prevalence/incidence) given the mean PGS at each percentile under the additive expectation (null model without PGSxE). Assuming the underlying risk distribution of overall population has a mean (μ) zero and variance (σ) 1, the liability threshold (*t*) can be determined from the prevalence in the overall population. Then, the mean of the underlying liability distribution (μ*_i_*) at each PGS percentile *i*, given the environmental effects are additive can be estimated:

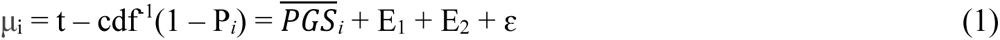

where, *t* is liability threshold determined from the overall population prevalence, P*_i_* is the observed prevalence at PGS percentile *i*, 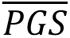 is the mean PGS at percentile *i*, E_1_ is the first exposure (0/1) and E_2_ is the second exposure (0/1). Then, the expected prevalence (P*i*’) at each PGS percentile *i* is computed as (assuming the variance of the underlying liability distribution (σ_i_) at each PGS percentile *i* is 1 under the null model):

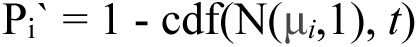

We performed 50 such iterations where u*_i_* was sampled from a distribution with variance equivalent to the residual error of the fitted model from equation (1). Similarly, the expected prevalence at each PGS percentile *i* can be computed by estimating the u*_i_* for the interaction model (including the PGSxE interaction terms) as:

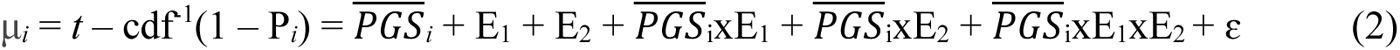

The expected disease risk at each PGS percentile under additive vs interaction models are depicted by grey dashed-curves and overlaid on the observed curves as illustrated in Figure 2 and Supplementary Figure S2 showing independent replication. We expect that deviations at the extremes will usually be associated with PGS×E influences on risk throughout the distribution. So, we computed the deviations of disease risk at the extremes of PGS (> or < +/-2 σ) between high vs low-risk exposures, denoted as delta^13^, in the observed and expected risk-PGS curves:

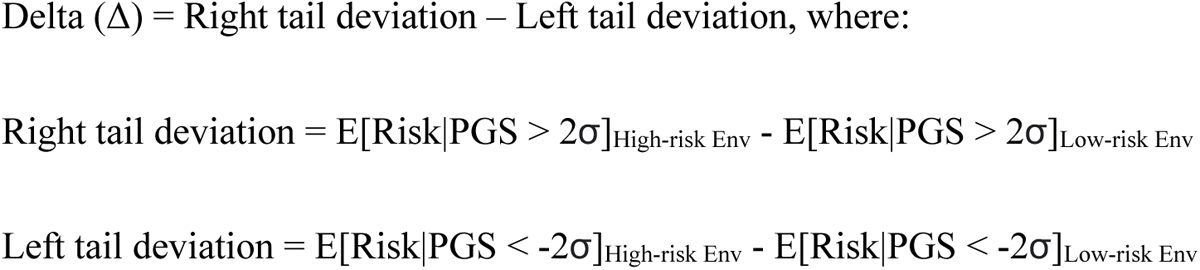

Then, the departure of Delta_observed_ from Delta_additive_ was computed as:

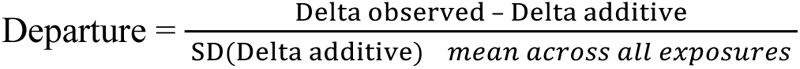

Combinations of exposures showing significant PGSxE were then identified if the departure of Delta_observed_ from Delta_additive_ was greater than 2 standard deviation units (sdu), illustrated in the chord diagrams (Figure 3 and Supplementary Figure S5). Of the 24,198 models evaluated across all diseases, 3742 (15.46%) exhibited a departure value exceeding 2 sdu, 3.1-fold more than expected by chance. Finally, we considered exposure-pairs that were deemed significant by both aforementioned models, resulting in a total of 746 models, which represents 3.1% of the total models assessed.

We further also estimated the excess disease variance explained by interaction effects from the overall liability threshold disease risk modeling i.e. variance explained by model including interactions terms (equation (2)) over additive model that does not include interaction term (equation (1)):

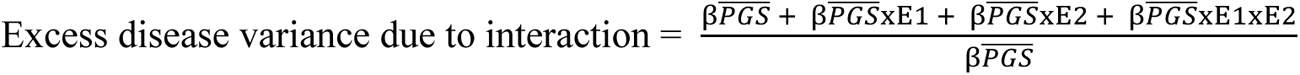

Supplementary Table 3 contains the metrics of the PGSxE models across significant exposures for each disease i.e., departure at PGS_extremes_, delta_observed_, delta_expected_ as well as the excess disease variance due to interaction effects from the overall model.

### Exposure-specific PGS analysis

To ensure that the deviations in risk across the PGS spectrum with respect to environmental exposures are not solely driven by prevalence/incidence differences or biased by PGS developed from GWAS of the overall population rather than exposure specific GWAS, we performed a series of sensitivity analyses. We considered sex as an exposure, owing to its large sample size, to construct exposure specific PGS (PGS_Exposure-specific_) for CAD in the UK Biobank. By splitting the males and females into 70% for GWAS (males: n=94,208, females: n=106,849) and 30% for PGS (males: n=62,805, females: n=71,244), matching the prevalence of CAD to the overall prevalence of males (16.5%) and females in the population (7.44%), we generated 10 such permutation sets. The GWAS models were adjusted for age and five genetic principal components. The exposure-specific log of odds ratio was divided by the standard error per variant to account for estimation noise and then used to construct exposure-specific PGS using the –score function in *PLINK*^58^. As a negative control, we also constructed exposure-specific PGS for randomly partitioned exposure groups (PGS_Exposure-specific (random partition)_), E_1rand_ and E_2rand_, keeping the sample size and prevalence the same as that in males and females. We then compared the exposure-specific estimates of effect sizes and PGS with the estimates from the overall population vs random environment. Specifically, spearman rank correlation was computed to evaluate if genetic effect sizes or PGS in specific exposures show higher or lesser correlation with estimates from the overall population to evaluate context-dependency. The spearman rank correlation also indicates whether the rank order of individuals is preserved when using PGS_Overall_ vs PGS_Exposure-specific._ Further, we also compared the shapes of risk-PGS curves with respect to males vs females using PGS_Overall_, PGS_Exposure-specific_ (exposure specific PGS constructed using exposure specific genetic effects in males and females separately) and PGS_Exposure-specific (random partition)_ for the randomly partitioned environment. Specifically, the deviations in CAD risk at median and top percentile of PGS were evaluated.

Supplementary Figure S3 illustrates this analysis. Notably, for both effect sizes (Supplementary Figure S3A) and PGSs (Supplementary Figure S3B), the correlation of exposure-specific estimates vs estimates from the overall population were higher for males (spearman rank rho_beta_ = 0.78 +/-0.001, rho_PRS_ = 0.9 +/-0.0008) compared to females (rho_beta_ = 0.64 +/-0.0023, rho_PRS_ = 0.73 +/-0.0025), indicating context-dependency. The prevalence-PGS curves generated with PGS_Overall_ (Supplementary Figure S3C) yielded higher prevalence in the top percentile for both males and females (males: 61.12% +/-0.75, females: 28.62% +/-0.37) indicating higher predictive ability of PGS_Overall_ to distinguish CAD cases vs controls than PGS_expspec_ (males: 52.75% +/-0.44, females: 19.8% +/-0.37, Supplementary Figure S3D). However, the deviations between prevalence-PGS curves with respect to males vs females was observed to be slightly higher when using PGS_Exposure-specific_ than PGS_Overall_, suggesting that the deviations of risk between exposures is slightly more conservative with PGS_Overall_ than PGS_Exposure-specific_. Whereas for the random environment partition, the correlations of genetic effect sizes (Supplementary Figure S3A) and PGS (Supplementary Figure S3B) between exposure-specific estimates vs estimates from overall population were similar for E_1rand_ and E_2rand_ and intermediate between those of males and females (E_1rand_: rho_beta_ = 0.76 +/-0.0018, rho_PRS_ = 0.86 +/-0.0016; E_2rand_: rho_beta_ = 0.7 +/-0.0019, rho_PRS_ = 0.8 +/-0.0013). The prevalence in the top percentile for random environment was the lower than that of males for E_1rand_ and higher than that of females for E_2rand_, (E_1rand_: 48.75% +/-0.44, E_2rand_: 24.38% +/-0.32), thus smaller deviations of disease risk with respect to random environment partitions (Supplementary Figure S3E).

The estimation of the expected deviation between environments is complicated for case-control traits because under the threshold liability model, odds ratios are a function of prevalence. Thus, to evaluate if the deviations in disease risk is not solely driven by CAD prevalence difference between males and females, we also estimated the exposure specific genetic effects and PGS_Exposure-specific_, keeping the sample size and prevalence of males and females equivalent to prevalence of CAD in the overall population of 11.7% and repeated the same analyses. Supplementary Figure S4 illustrates the analyses with prevalence-matched exposure groups.

### Predictive accuracy of PGS models

We evaluated the predictive accuracy of PGS by estimating the Nagelkerke R^2^ from the logistic regression of four models:

i. Overall population, no environmental stratification: Disease ∼ PGS + covariates;
ii. In low-risk exposures (E_00_): Disease ∼ PGS + covariates;
iii. In high-risk exposure (E_11_): Disease ∼ PGS + covariates;
iv. Composite exposure model with interaction: Disease ∼ PGS + E_1_ + E_2_ + PGSxE_1_ + PGSxE_2_ and PGSxE_1_xE_2_ + covariates.

The covariates included age and five genetic principal components. The R^2^ was estimated using the ‘lrm’ function of ‘rms’ package in R (http://cran.r-project.org/web/packages/rms/). Across major exposures exhibiting multiple interactions for each disease, the average R^2^ value was computed across its combinations as shown in Figure 4 and Supplementary Figure S13.

### Amplification of genetic effects

We estimated the genetic effect sizes in high (E_11_) vs low-risk (E_00_) exposures for major exposures and its combinations identified for each disease. The exposure-specific genetic effects were estimated by running GWAS in the UKB for a subset of independent SNPs that were used for PGS construction for each disease (Supplementary Table S1), adjusting for age and genetic principal components. We next used multivariate adaptive shrinkage (mash)^47^ to examine the mixture of covariance relationships (correlation and difference in magnitude) of SNP effect sizes along with their estimation noise (standard error), between high-risk exposures (H: E_11_) vs low-risk exposures (L: E_00_). Mash learns from the data by estimating mixture proportions of various pre-defined covariance matrices representing different patterns of effects. It assigns weights to covariance matrices using maximum likelihood, assigning low weight to matrices that capture fewer patterns and high weight to those which capture more patterns. We utilized the 66 pre-defined covariance matrices from Zhu, et al^20^ used to study the amplification of genetic effects. These included correlation values of -1, -0.75, -0.5, -0.25, 0, 0.25, 0.5, 0.75, 1 and variance (denoting the difference in magnitude of effect sizes) being equal in both high-risk (H i.e. E_11_) and low-risk (L i.e. E_00_) exposure, L-specific or higher in low-risk exposure i.e. L 1.5x, L 2x, L3x and H-specific or higher in high-risk exposure i.e. H 1.5x, H 2x, H 3x. The remainder weight is assigned to the no-effect matrix. Essentially, mash quantifies the proportion of SNPs that follow each specific pattern of magnitude and correlation between high vs low-risk exposures. Supplementary Figure S14 illustrates the amplification of genetic effects in past smokers with low omega 6 fatty acids compared to non-smokers with high omega 6 levels. The majority of the genetic effect sizes are well correlated between high vs low-risk exposures (weight assigned to covariance matrix with correlation of 1) of which 39.36% are equal in magnitude, 55.07% are 2x higher in high-risk exposure and 2.32% are 3x higher in high-risk exposure i.e. past smokers with reduced omega 6 levels. 3.25% SNPs are assigned correlation of 0.5 and are 2x higher in high-risk exposure, while a small proportion 0.0006% SNPs show no correlation but are specific to low-risk exposure. The net amplification effect is computed as the percentage of variants with genetic effect sizes higher in the high-risk exposure compared to low-risk exposure (H > L) minus percentage of variants with genetic effect sizes higher in low-risk exposure compared to high-risk exposure (L < H). The net amplification of genetic effects in past smokers with reduced omega-6 levels for CAD is 60.62% H>L (Supplementary Figure S14A). Thus, the genetic effect sizes between high vs low-risk exposure are very highly correlated but amplified in magnitude in the high-risk exposure. This finding is consistent across the majority of exposure combinations showing PGSxE effects, as shown in Figure 5 and Supplementary Figure S15. This implies that amplification (rather than uncorrelated effects) is the mechanism of PGSxE leading to differences in genetic variance between exposures and thus excess disease risk in high-risk exposure. SNP-heritability was also computed in low and high-risk exposures to compare the difference in genetic variance using BOLT-REML^60^. Supplementary Table S4 contains the net amplification effect (%) across significant exposures showing PGSxE and diseases.

### Proportion needed to benefit

Building on the idea of the number needed to treat (NNT)^6,7,49^, the proportion needed to benefit is defined as a measure of how many people must alter their behavior (based on modifiable exposures such as diet or lifestyle) in order to benefit one person. It is computed as the inverse of the difference between area under risk-PGS curves (AUC) for the high-risk exposure (E_11_) minus that for the low-risk exposure (E_00_) at different PGS thresholds, scaled by the proportion of population sampled at each threshold.

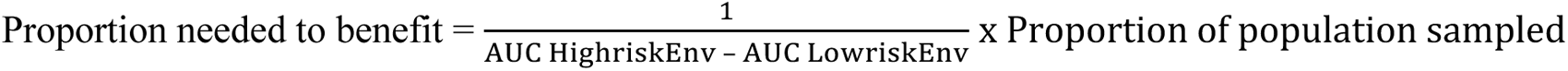

We computed the PNB in the overall population i.e. PGS percentile > 0 (overall population, without consideration of genetics) and increasing PGS thresholds i.e. PGS percentile > 10, PGS percentile > 20 and so on, up to PGS percentile > 80 (top 20% of the population) and PGS percentile > 90 (top 10% of the population). The gain in benefit was computed as the ratio of proportion of the population who might benefit from lifestyle intervention at a given PGS threshold over the proportion of the overall population who would benefit without consideration of genetics (i.e. PGS percentile > 0). Further, we also computed the precision (positive predictive values), negative predictive values, sensitivity and specificity at each PGS threshold in high vs low risk exposures. The definitions for computing these metrics are provided in Supplementary Table S5 illustrating an example of proportion needed to benefit for a modifiable exposure, bread type and smoking status with respect to CAD risk and increasing PGS thresholds.

## Data Availability

UK Biobank individual-level genotype and phenotype data are available through application at http://www.ukbiobank.ac.uk. The results to explore risk curves across all exposures and diseases will be made available through an R shiny application upon publication.

## Code Availability

Code will be available upon publication.

## Acknowledgments

We thank Arbel Harpak, Alison Motsinger-Reif, and Raghav Tandon for their insightful discussions and feedback. We thank the study participants of the UK Biobank. This work was supported by a grant from the U.S. National Institutes of Health to G.G. (R01-DK119991). Analyses using the UK Biobank Resource were performed under Institutional Review Board–approved application number 17984.

## Supplementary Material

**Figure S1:**
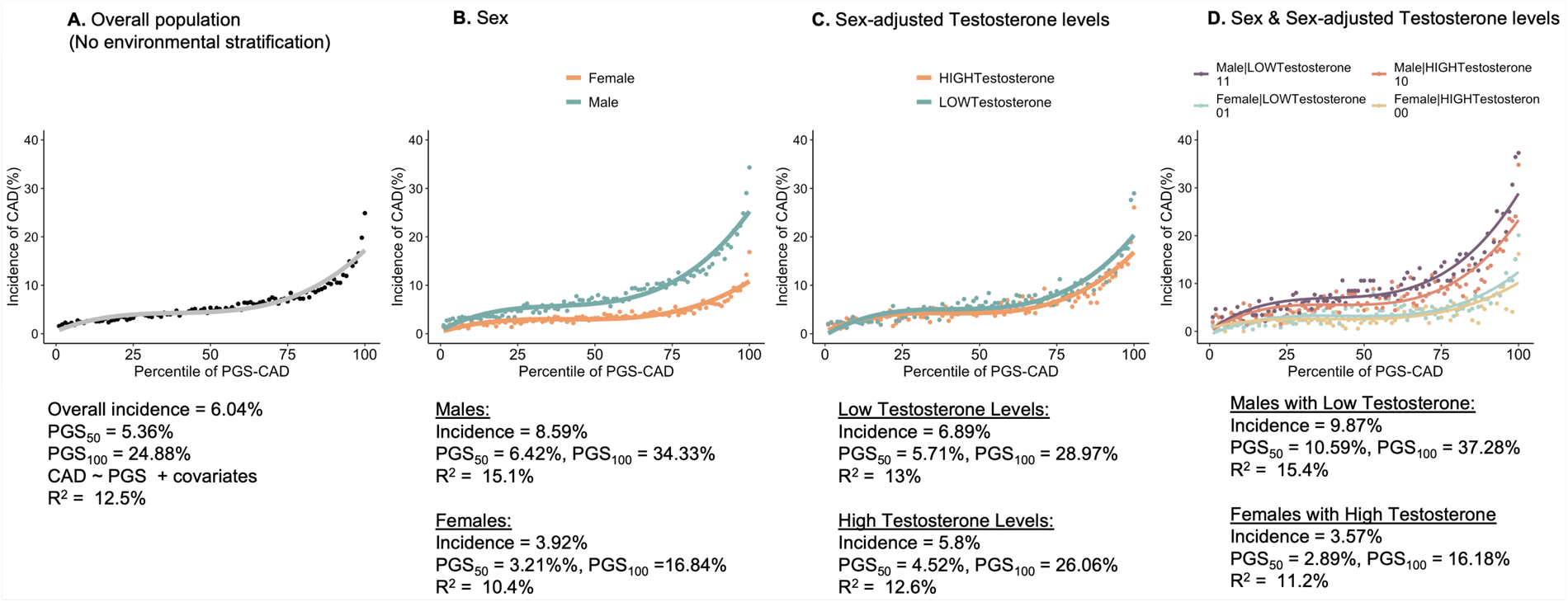
Incidence of CAD vs percentile of PGS curves for (A) Overall population. (without any environmental stratification) has CAD incidence 6.04%, incidence at 50^th^ percentile of PGS is 5.36% whereas incidence in the top percentile of PGS is 24.88%. The predictive accuracy of PGS, R^2^ is 12.5%. **(B) Stratified by sex**: Males have an overall incidence of 8.59%, incidence at 50^th^ percentile of PGS is 6.42% whereas incidence in the top percentile of PGS is 34.33%. The predictive accuracy of PGS, R^2^ is 15.1%. Females have an overall incidence 3.92%, incidence at 50^th^ percentile of PGS is 3.21% whereas incidence in the top percentile of PGS is 16.84%. The predictive accuracy of PGS, R^2^ is 10.4%. **(C) Stratified by sex-adjusted testosterone levels**: Low testosterone group has an overall incidence of 6.89%, incidence at 50^th^ percentile of PGS is 5.71% whereas incidence in the top percentile of PGS is 28.97%. The predictive accuracy of PGS, R^2^ is 13%. High testosterone group has an overall incidence 5.8%, incidence at 50^th^ percentile of PGS is 4.52% whereas incidence in the top percentile of PGS is 26.06%. The predictive accuracy of PGS, R^2^ is 12.6%. **(D) Stratified by combination of exposures – sex and sex-adjusted testosterone levels**: Males with low testosterone levels (E_11_: purple curve) have an overall incidence of 9.87%, incidence at 50^th^ percentile of PGS is 10.59% whereas incidence in the top percentile of PGS is 37.28%. The predictive accuracy of PGS, R^2^ is 15.4%. Females with high testosterone levels (E_00_: yellow curve) have an overall incidence 3.57%, incidence at 50^th^ percentile of PGS is 2.89% whereas incidence in the top percentile of PGS is 16.18%. The predictive accuracy of PGS, R^2^ is 11.2%.

**Figure S2:**
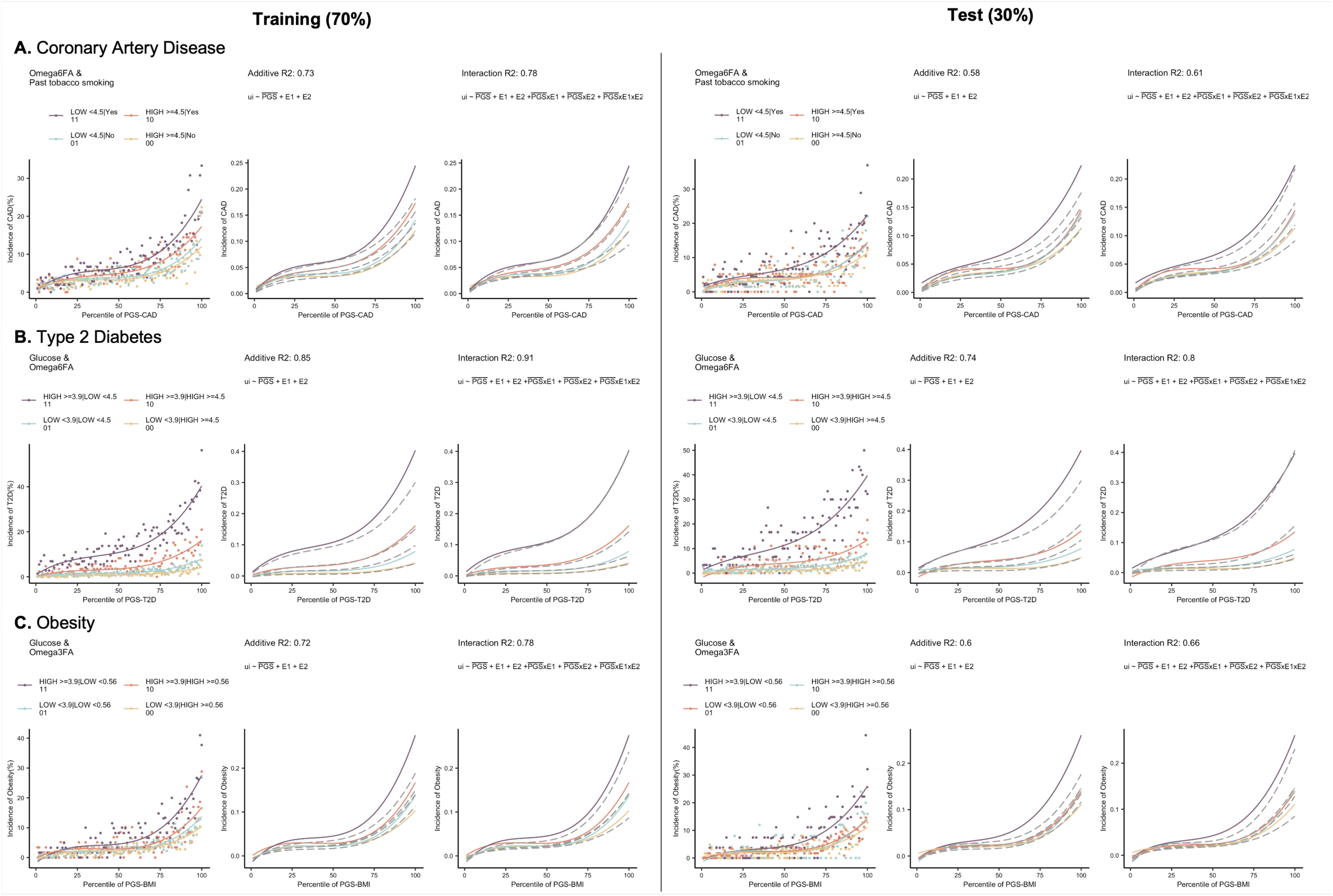
Assessment of interactions between PGS and exposures shown on 70% training set (left) and 30% independent test set (right). **(A)** Incidence of CAD vs percentile PGS-CAD with respect to omega 6 fatty acids and past tobacco smoking, **(B)** Incidence of T2D vs percentile of PGS-T2D with respect to glucose and omega 6 fatty acid levels, **(C)** Incidence of obesity vs percentile of PGS-BMI with respect to glucose and omega 3 fatty acid levels. The second panel in each case shows the expected risk if the environmental effects are additive as shown by the grey dashed curves obtained using liability threshold disease risk modeling, overlaid on the observed curves. The third panel shows the expected risk (grey dashed curves) under the interaction model.

**Figure S3:**
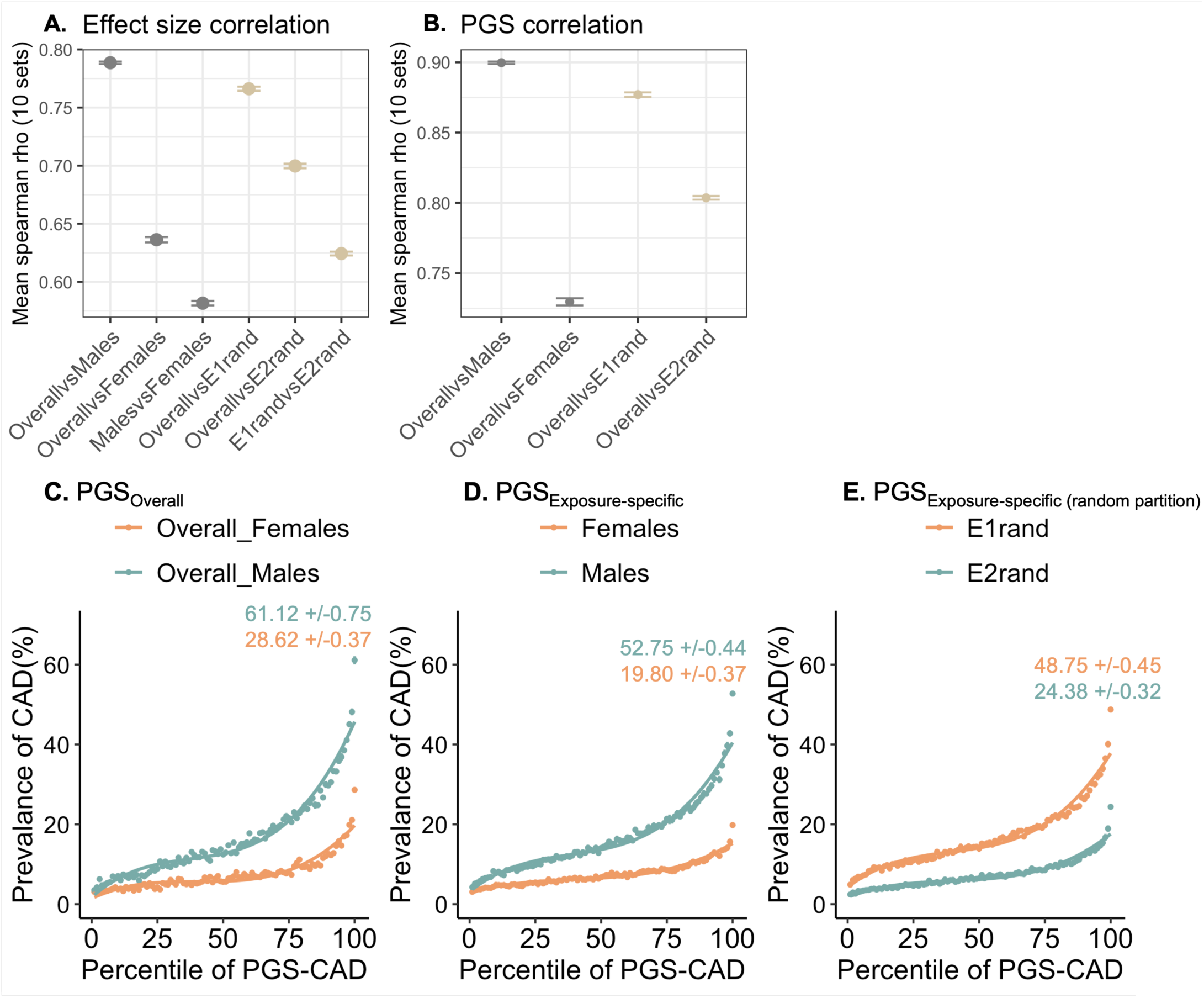
Exposure specific GWAS and PGS analyses considering sex as an exposure vs CAD. 10 permutation sets were generated, where 70% samples were retained for GWAS and 30% for PGS, and prevalence of CAD in males is 16.5% and females is 7.44% equivalent to that observed in the population. **(A)** Spearman rank correlation of effect size between estimates from overall population and exposure specific estimates. **(B)** Spearman rank correlation of PGS between estimates from overall population and exposure specific estimates **(C)** Prevalence of CAD vs percentile PGS-CAD with respect to males vs females, where PGS is computed from GWAS of overall population (PGS_overall_). **(D)** Prevalence of CAD vs percentile PGS-CAD with respect to males vs females, where PGS is computed from exposure specific GWAS in males and females respectively (PGS_Exposure-specific_). **(E)** Prevalence of CAD vs percentile PGS-CAD with respect to males vs females, where PGS is computed from exposure specific GWAS in the randomly partitioned environments (PGS_Exposure-specific (random partition)_).

**Figure S4:**
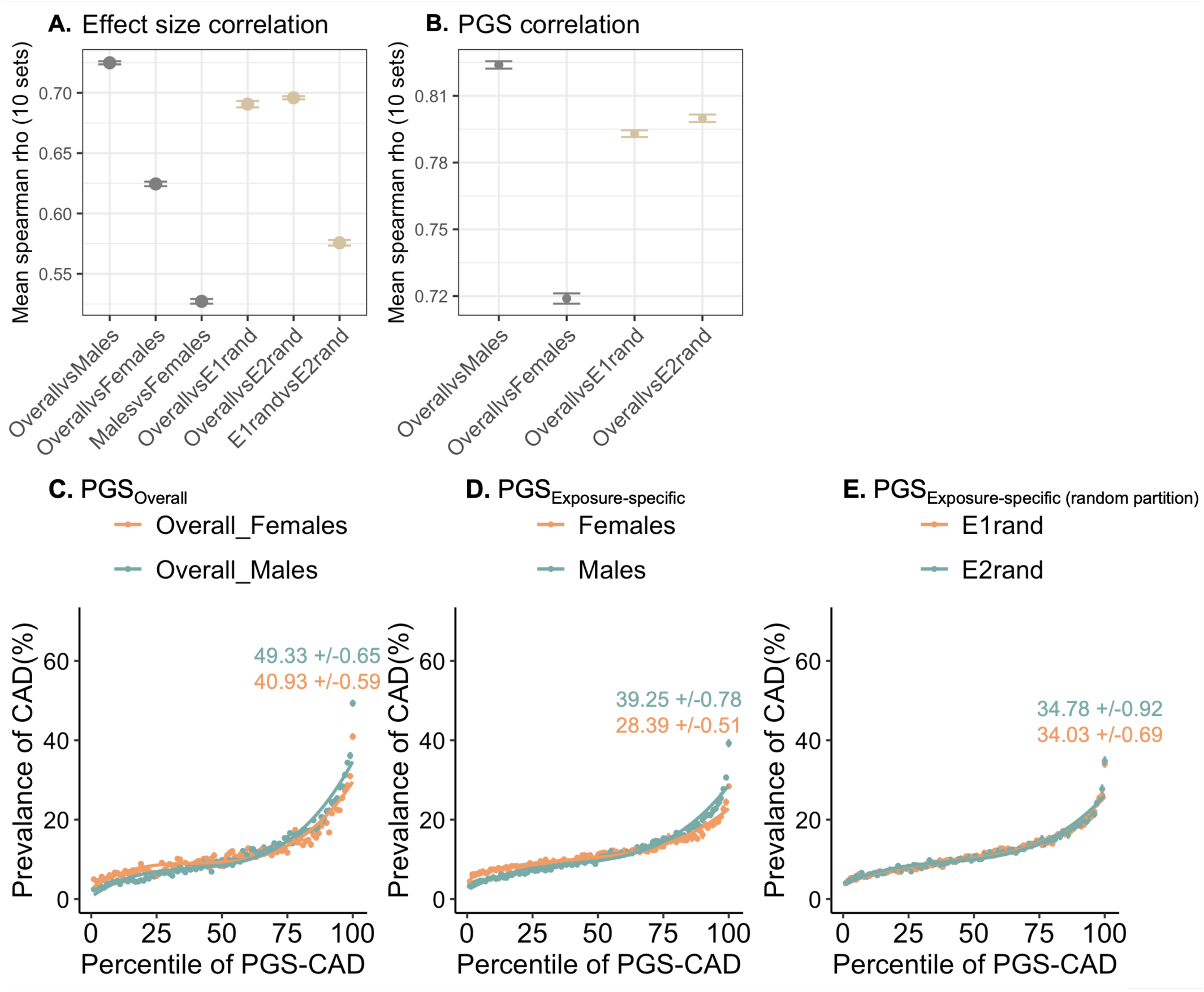
Prevalence & sample size matched exposure specific GWAS and PGS analyses considering sex as an exposure vs CAD. 10 permutation sets were generated, where 70% samples were retained for GWAS and 30% for PGS. The prevalence of CAD in males and females is equivalent to that in the overall prevalence i.e. 11.7%. **(A)** Spearman rank correlation of effect sizes between estimates from overall population and exposure specific estimates. **(B)** Spearman rank correlation of PGS between estimates from overall population and exposure specific estimates **(C)** Prevalence of CAD vs percentile PGS-CAD with respect to males vs females, where PGS is computed from GWAS of overall population (PGS_overall_). **(D)** Prevalence of CAD vs percentile PGS-CAD with respect to males vs females, where PGS is computed from exposure specific GWAS in males and females respectively (PGS_Exposure-specific_). **(E)** Prevalence of CAD vs percentile PGS-CAD with respect to males vs females, where PGS is computed from exposure specific GWAS in the randomly partitioned environments (PGS_Exposure-specific (random partition)_).

**Figure S5:**
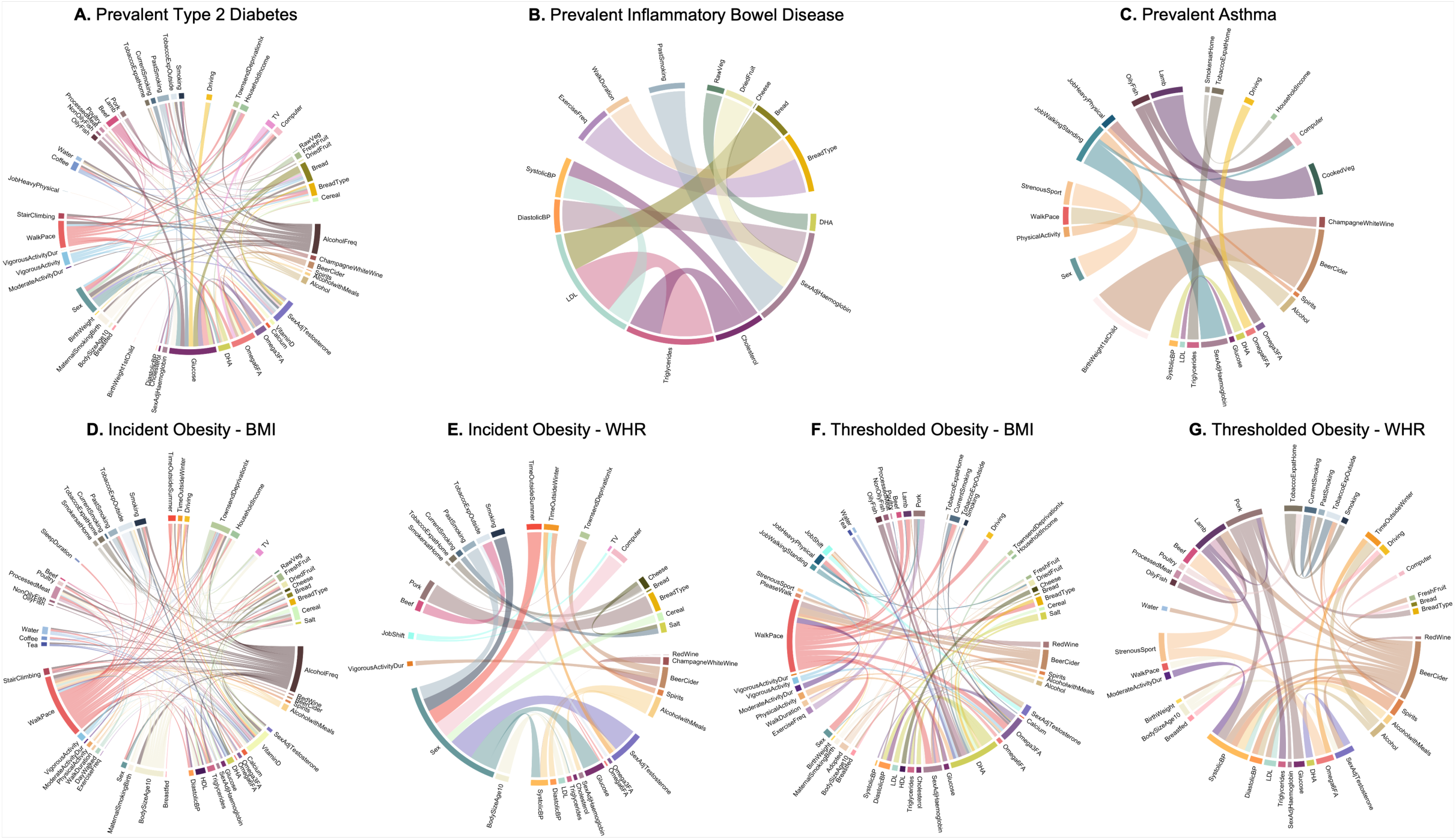
Combinations of exposures showing significant PGSxE for common diseases. **(A)** Prevalent type 2 diabetes (T2D), **(B)** Prevalence inflammatory bowel disease (IBD), **(C)** Prevalent Asthma, **(D)** Incident obesity assessed with respect to PGS-BMI, **(E)** Incident obesity assessed with respect to PGS-WHR, **(F)** Thresholded obesity – BMI, **(G)** Thresholded obesity – waist-to-hip ratio (WHR). The width of the arc denotes the delta_observed_ i.e. deviations of disease risk at the extremes of PGS between high-vs low-risk exposure and the number of linkages per node denote the multiple interactions exhibited by key or major exposures for each disease.

**Figure S6:**
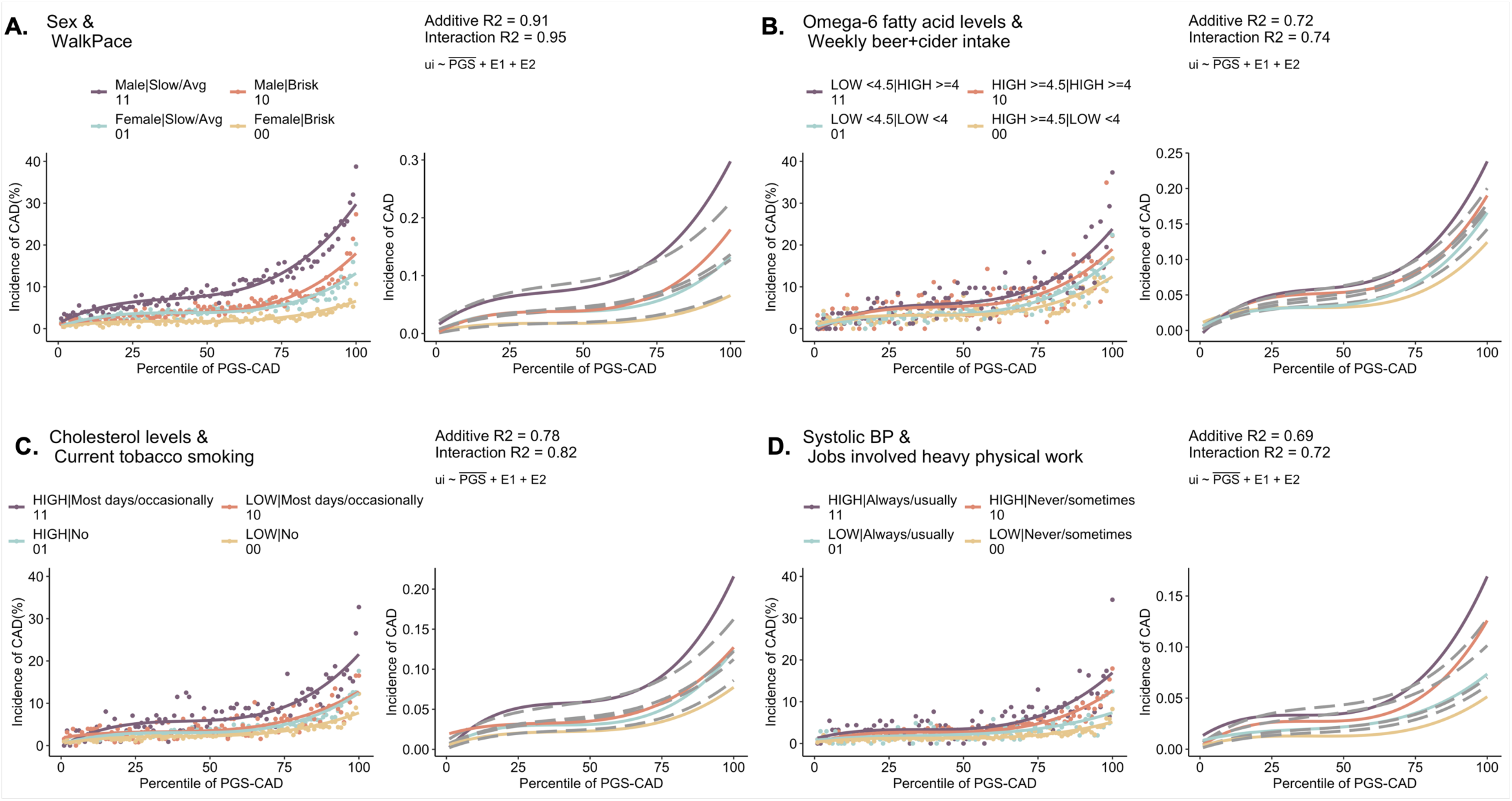
Notable PGSxE cases for incident CAD. The curves show incidence of CAD vs percentile PGS-CAD with respect to combinations of environmental exposures: **(A)** Sex and walk pace, **(B)** Omega 6 fatty acid levels and average weekly beer or cider intake, **(C)** Cholesterol levels and current tobacco smoking, **(D)** Systolic blood pressure and jobs involved heavy physical work.

**Figure S7:**
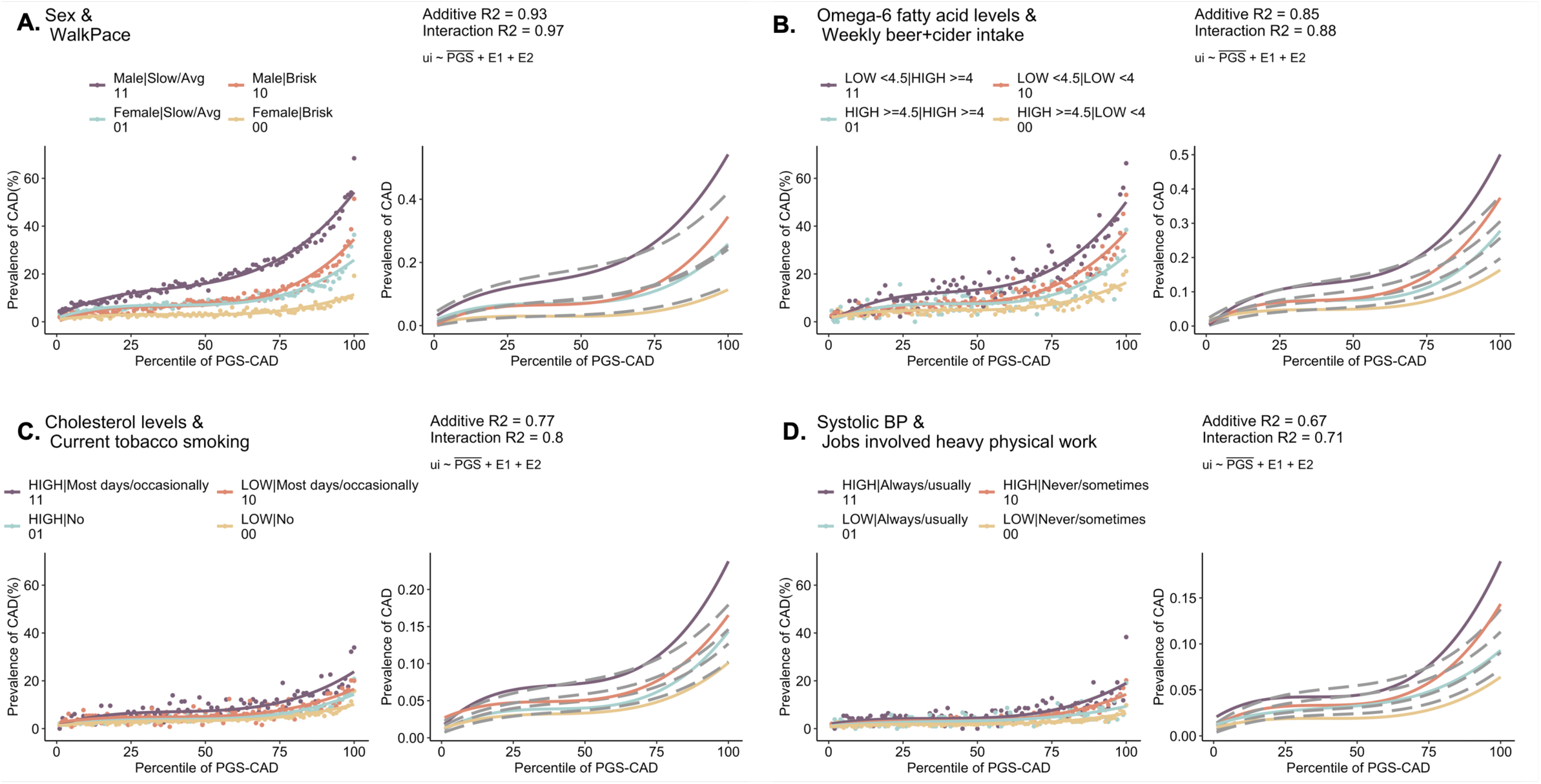
Notable PGSxE cases for prevalent CAD. The curves show prevalence of CAD vs percentile PGS-CAD with respect to combinations of environmental exposures: **(A)** Sex and walk pace, **(B)** Omega 6 fatty acid levels and average weekly beer or cider intake, **(C)** Cholesterol levels and current tobacco smoking, **(D)** Systolic blood pressure and jobs involved heavy physical work.

**Figure S8:**
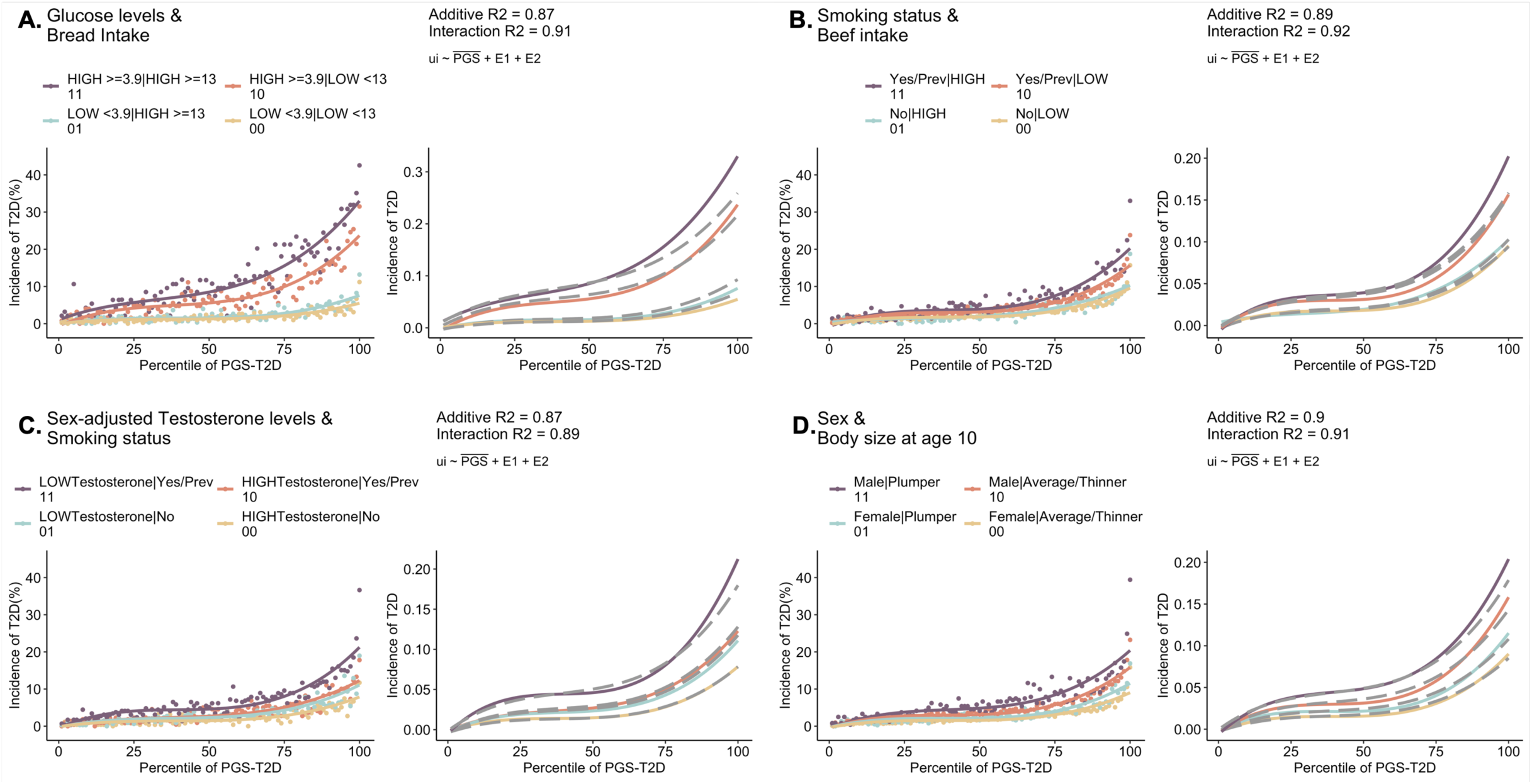
Notable PGSxE cases for incident T2D. The curves show incidence of T2D vs percentile PGS-T2D with respect to combinations of environmental exposures: **(A)** Glucose levels and bread intake **(B)** Smoking status and beef intake, **(C)** Sex adjusted testosterone levels and smoking status, **(D)** Sex and body size at age 10.

**Figure S9:**
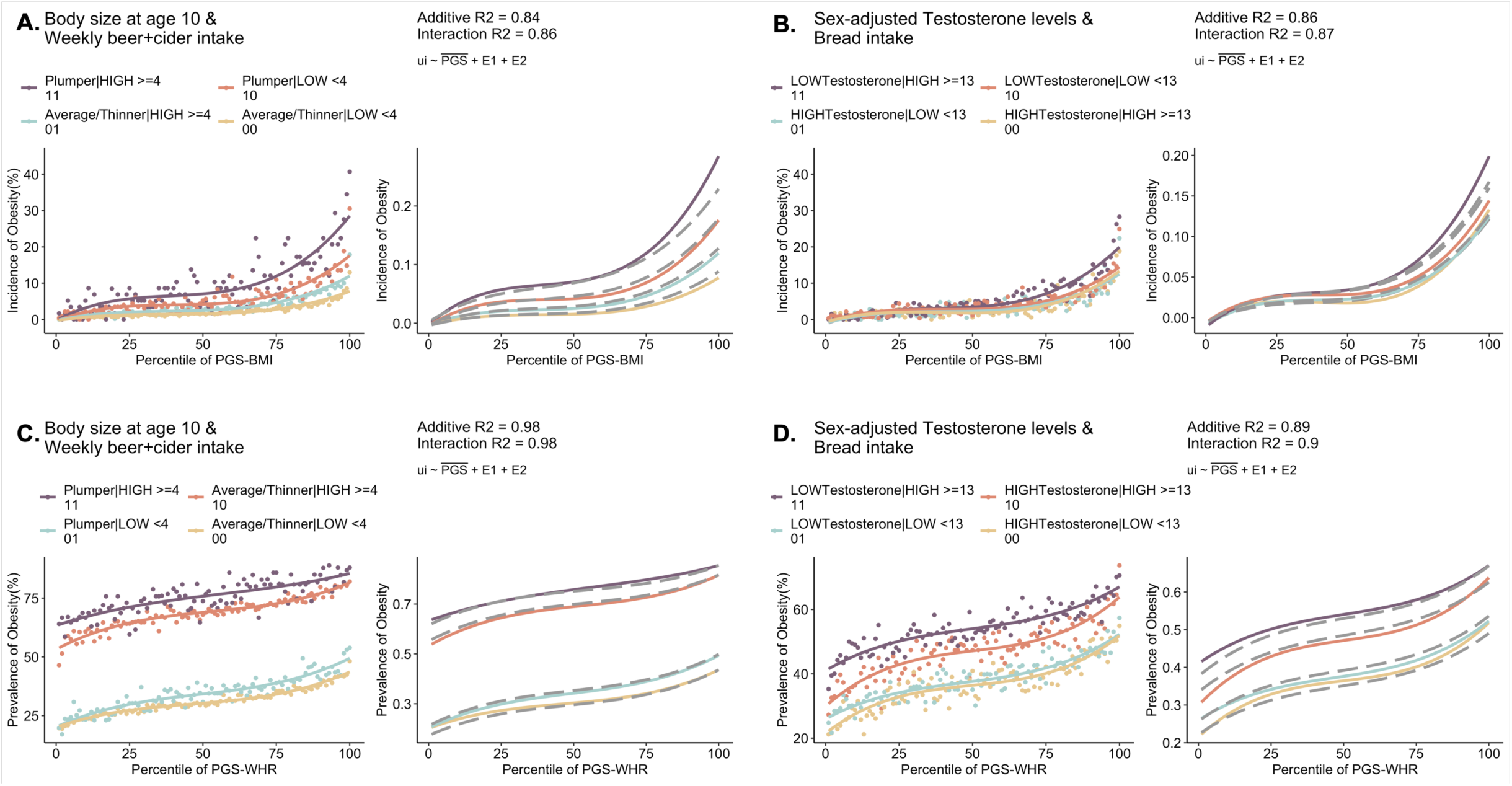
Notable cases of incident obesity vs PGS-BMI and prevalent obesity vs PGS-WHR with respect to (A, C) body size at age 10 and average weekly beer or cider intake, **(B, D)** sex-adjusted testosterone levels and bread intake, respectively.

**Figure S10:**
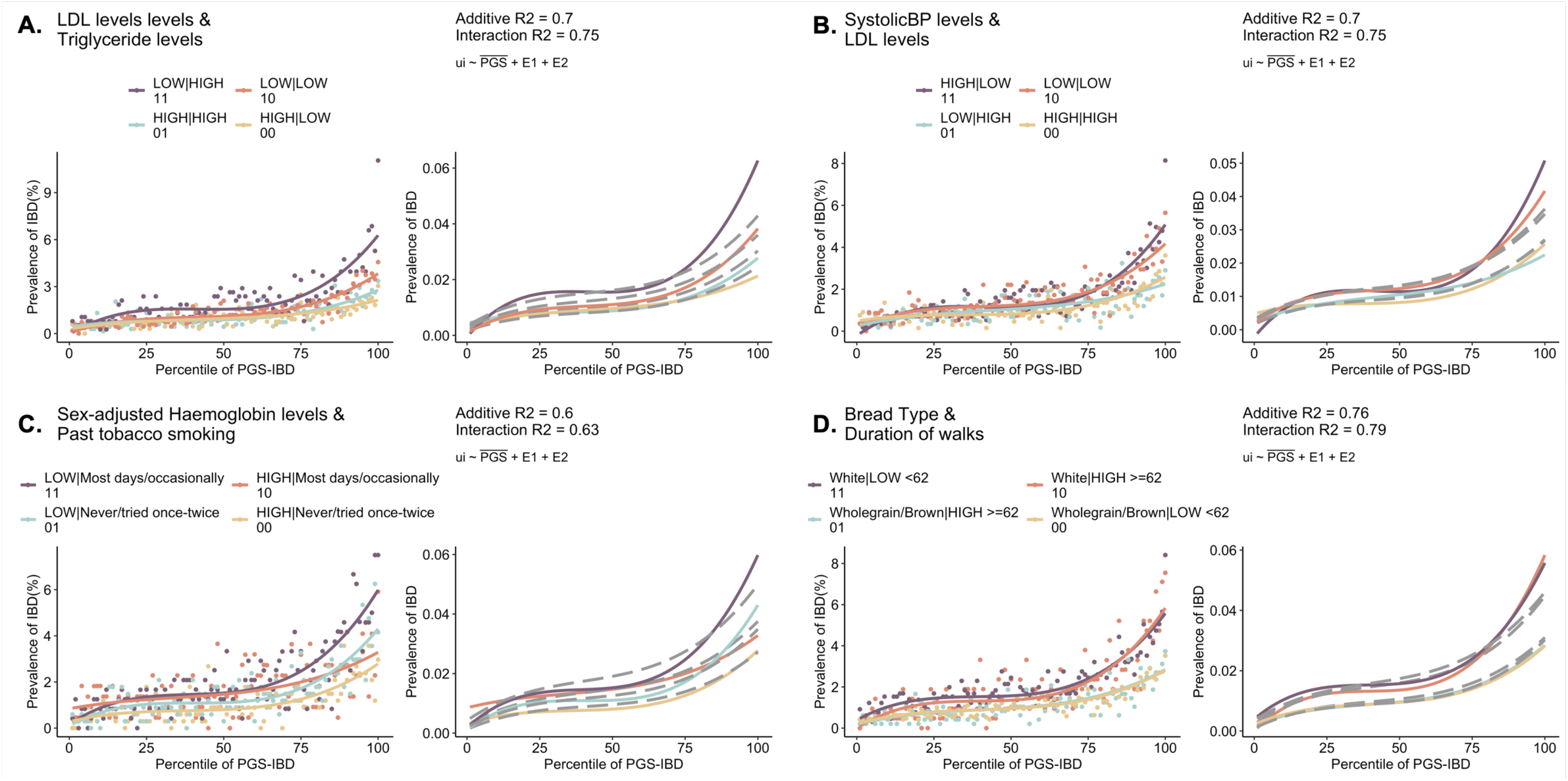
Notable PGSxE cases for prevalent IBD. The curves show prevalence of IBD vs percentile PGS-IBD with respect to combinations of environmental exposures: **(A)** LDL and triglyceride levels **(B)** Systolic blood pressure and LDL levels, **(C)** Sex adjusted hemoglobin concentration and past tobacco smoking status, **(D)** Bread type as white or wholegrain/brown and duration of walks.

**Figure S11:**
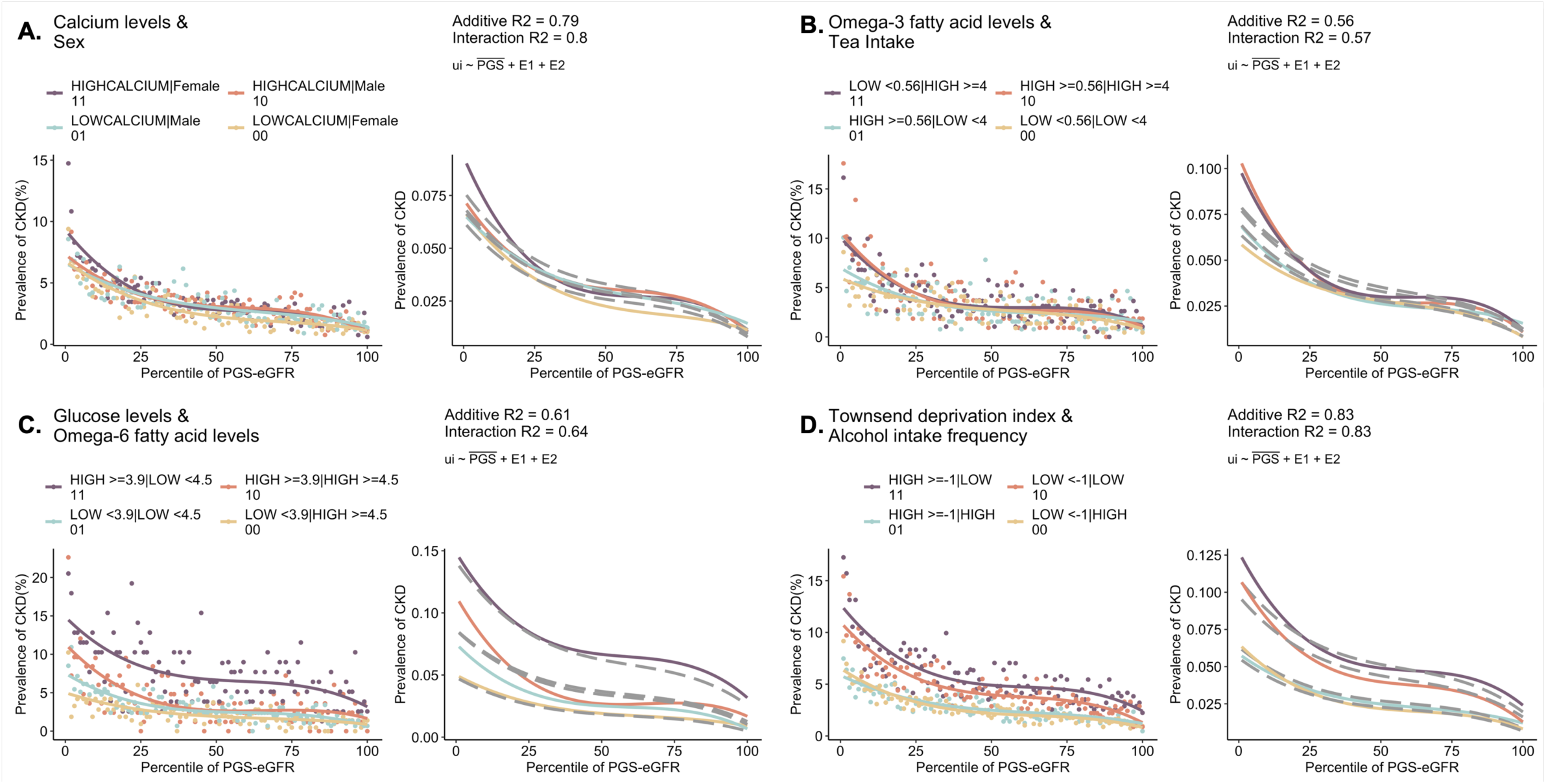
Notable PGSxE cases for prevalent CKD. The curves show prevalence of CKD vs percentile PGS-CKD with respect to combinations of environmental exposures: **(A)** Sex and calcium levels **(B)** omega 3 fatty acid levels and tea intake, **(C)** Glucose and omega 6 fatty acid levels, **(D)** Townsend deprivation index and alcohol intake frequency.

**Figure S12:**
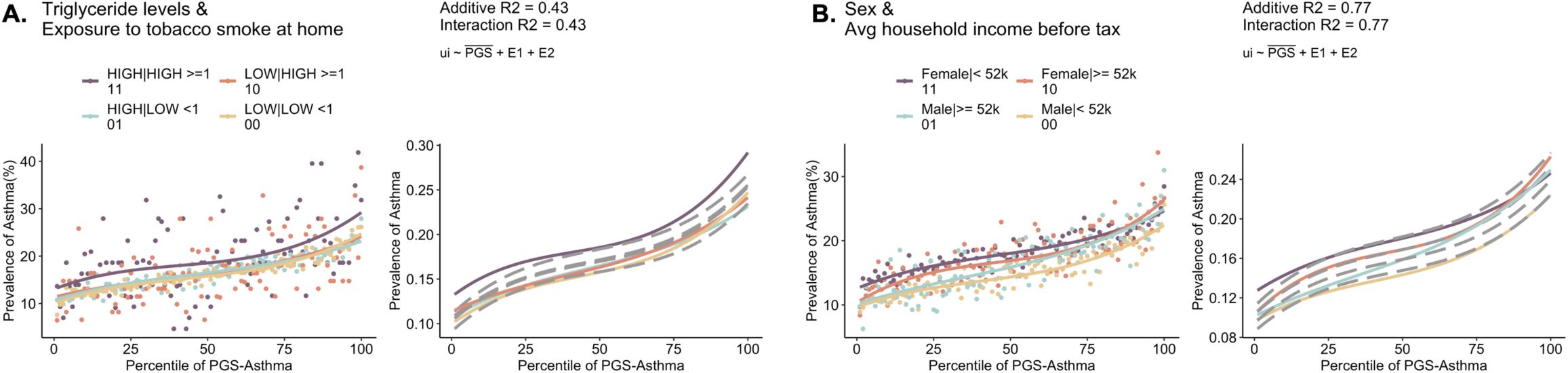
Notable PGSxE cases for prevalent Asthma. The curves show prevalence of Asthma vs percentile PGS-Asthma with respect to combinations of environmental exposures: **(A)** Triglyceride levels and exposure to tobacco smoke at home **(B)** Sex and average total household income before tax.

**Figure S13:**
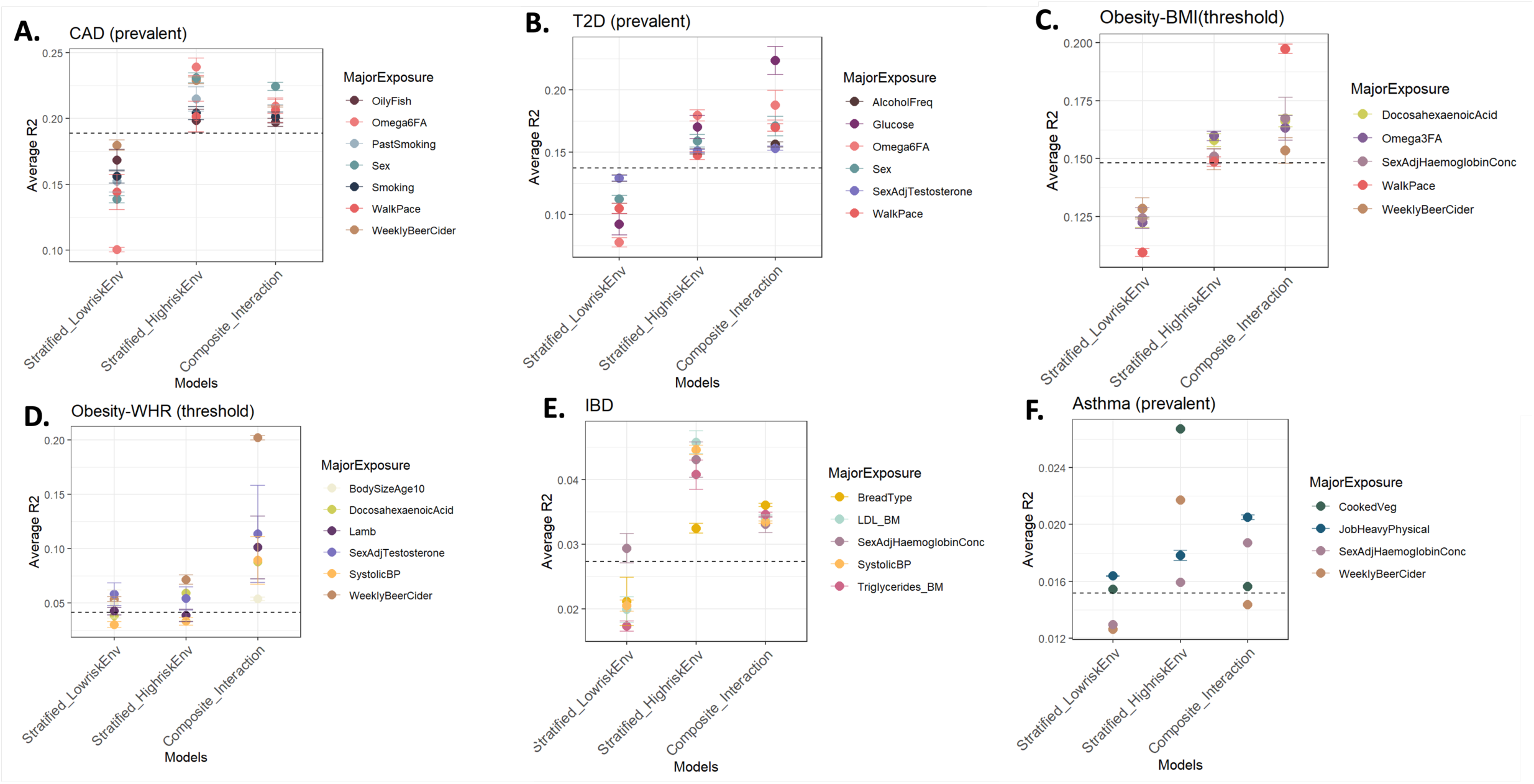
Gain in PGS prediction accuracy in high-risk exposures and composite exposure model including the interaction effects. The average R2 across significant exposure-pairs is shown for major exposures across each disease: **(A)** Prevalent CAD, **(B)** Prevalent T2D, **(C)** Thresholded Obesity-BMI, **(D)** Thresholded Obesity-WHR, **(E)** IBD, **(F)** Asthma.

**Figure S14:**
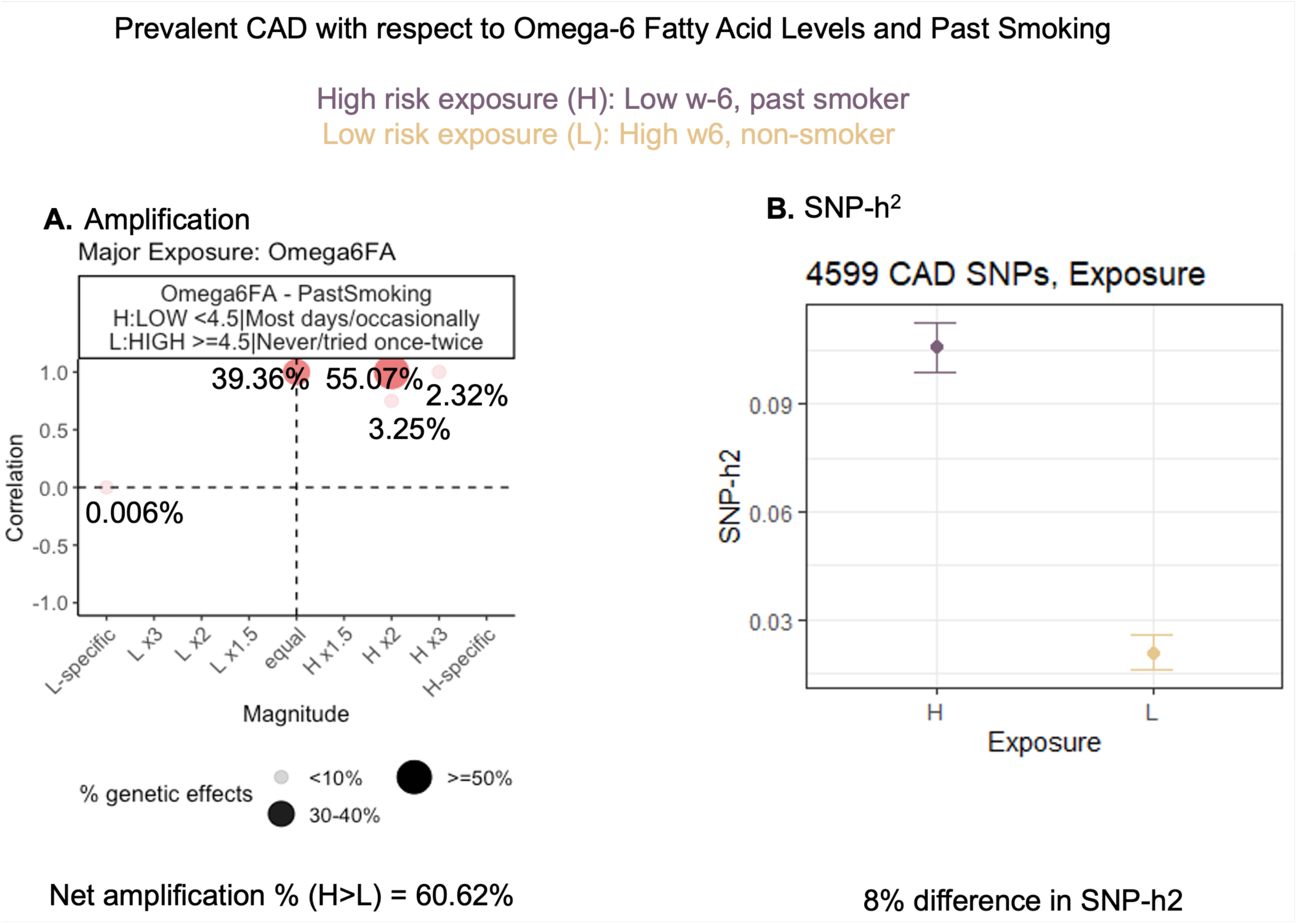
Amplification and SNP-heritability assessed for 4599 CAD SNPs with respect to omega 6 fatty acid levels and past tobacco smoking status. **(A)** Amplification of genetic effects in the high-risk exposure (H) i.e. low omega-6 fatty acid levels and past smoker compared to low-risk exposure (L) i.e. high omega-6 levels and non-smoker. The x-axis denotes the magnitude, whether H>L or L>H and the y-axis denotes the correlation values. **(B)** SNP-h2 computed using BOLT-REML in high-and low-risk exposures.

**Figure S15:**
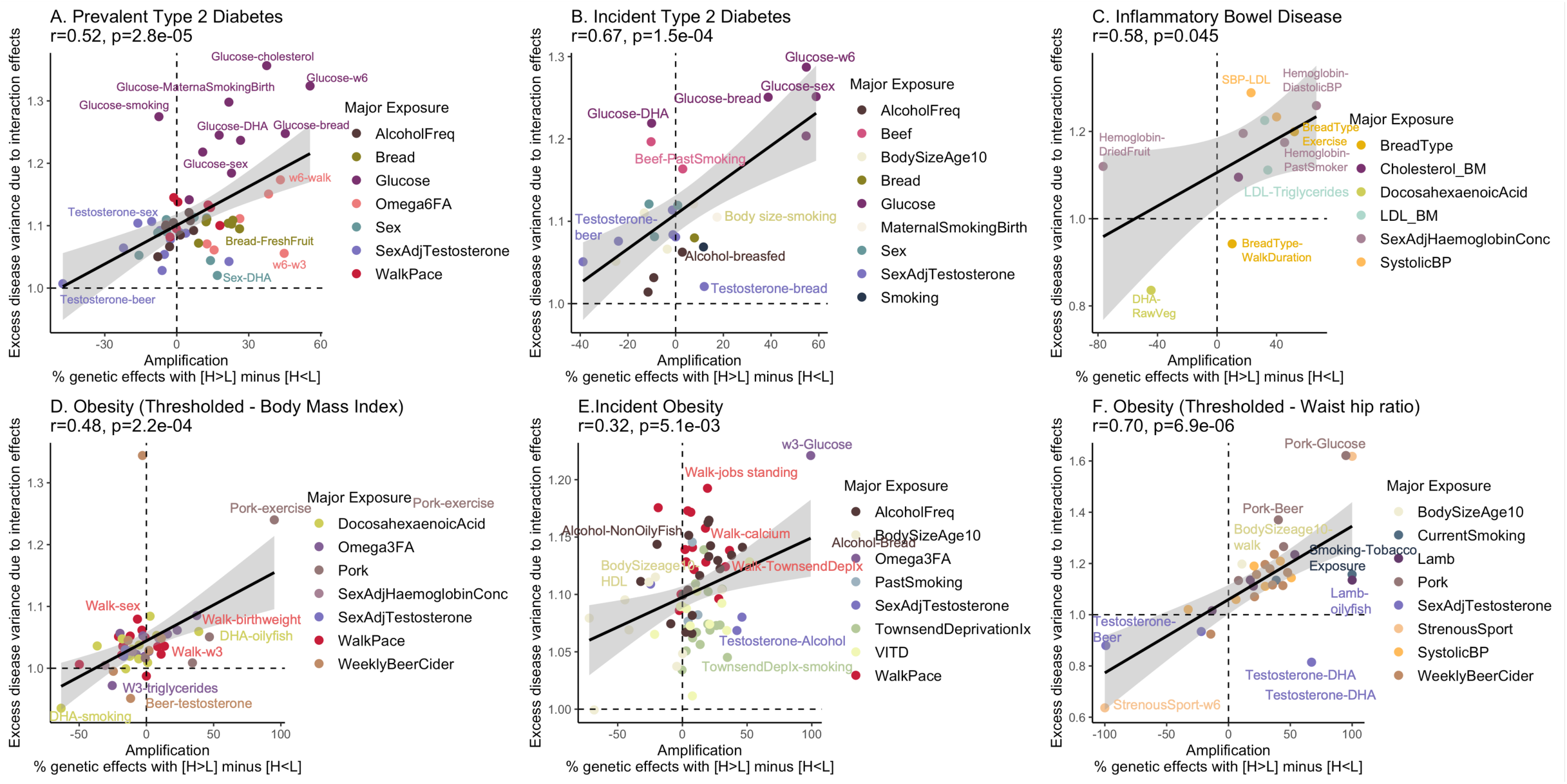
PGSxE shows a positive relationship with amplification of genetic effects. The x-axis denotes the net amplification effect computed as the percentage of genetic effects with H (E_11_) > L (E_00_) minus percentage of genetic effect with H (E_11_) < L (E_00_). The y-axis denotes the excess disease variance due to interaction effects computed from the liability threshold disease risk modeling. Shown exposure pairs for **(A)** Prevalent T2D, **(B)** Incident T2D, **(C)** Prevalent IBD, **(D)** Thresholded Obesity-BMI, **(E)** Incident obesity, **(F)** Thresholded Obesity-WHR.

## REFERENCES

1. Lewis, A.C.F. et al. Managing differential performance of polygenic risk scores across groups: Real-world experience of the eMERGE Network. Am J Hum Genet 111, 999–1005 (2024).

2. Lennon, N.J. et al. Selection, optimization and validation of ten chronic disease polygenic risk scores for clinical implementation in diverse US populations. Nature Medicine 30, 480–487 (2024).

3. Jermy, B. et al. A unified framework for estimating country-specific cumulative incidence for 18 diseases stratified by polygenic risk. Nature Communications 15, 5007 (2024).

4. Xiang, R. et al. Recent advances in polygenic scores: translation, equitability, methods and FAIR tools. Genome Medicine 16, 33 (2024).

5. Samani, N.J. et al. Polygenic risk score adds to a clinical risk score in the prediction of cardiovascular disease in a clinical setting. *European Heart Journal*, ehae342 (2024).

6. Gibson, G. Going to the negative: genomics for optimized medical prescription. Nat Rev Genet 20, 1–2 (2019).

7. Gibson, G. On the utilization of polygenic risk scores for therapeutic targeting. PLoS Genet 15, e1008060 (2019).

8. Martin, A.R. et al. Clinical use of current polygenic risk scores may exacerbate health disparities. Nat Genet 51, 584–591 (2019).

9. Wang, Y., Tsuo, K., Kanai, M., Neale, B.M. & Martin, A.R. Challenges and Opportunities for Developing More Generalizable Polygenic Risk Scores. Annu Rev Biomed Data Sci 5, 293–320 (2022).

10. Kachuri, L. et al. Principles and methods for transferring polygenic risk scores across global populations. Nature Reviews Genetics 25, 8–25 (2024).

11. Wang, Y. et al. Aspiring toward equitable benefits from genomic advances to individuals of ancestrally diverse backgrounds. Am J Hum Genet 111, 809–824 (2024).

12. Madden, E.B. et al. Advancing genomics to improve health equity. Nat Genet 56, 752–757 (2024).

13. Nagpal, S., Tandon, R. & Gibson, G. Canalization of the Polygenic Risk for Common Diseases and Traits in the UK Biobank Cohort. Mol Biol Evol 39(2022).

14. Durvasula, A. & Price, A.L. Distinct explanations underlie gene-environment interactions in the UK Biobank. medRxiv (2024).

15. Herrera-Luis, E., Benke, K., Volk, H., Ladd-Acosta, C. & Wojcik, G.L. Gene-environment interactions in human health. Nat Rev Genet (2024).

16. Virolainen, S.J., VonHandorf, A., Viel, K., Weirauch, M.T. & Kottyan, L.C. Gene-environment interactions and their impact on human health. Genes Immun 24, 1–11 (2023).

17. Marderstein, A.R. et al. Leveraging phenotypic variability to identify genetic interactions in human phenotypes. Am J Hum Genet 108, 49–67 (2021).

18. Mostafavi, H. et al. Variable prediction accuracy of polygenic scores within an ancestry group. Elife 9(2020).

19. Hui, D. et al. Risk factors affecting polygenic score performance across diverse cohorts. (Cold Spring Harbor Laboratory, 2023).

20. Zhu, C. et al. Amplification is the primary mode of gene-by-sex interaction in complex human traits. Cell Genom 3, 100297 (2023).

21. Hermisson, J., Hansen, T.F. & Wagner, G.P. Epistasis in polygenic traits and the evolution of genetic architecture under stabilizing selection. Am Nat 161, 708–34 (2003).

22. Wagner, G.P., Booth, G. & Bagheri-Chaichian, H. A Population Genetic Theory of Canalization. Evolution 51, 329–347 (1997).

23. Gibson, G. & Lacek, K.A. Canalization and Robustness in Human Genetics and Disease. Annu Rev Genet 54, 189–211 (2020).

24. Gibson, G. Decanalization and the origin of complex disease. Nature Reviews Genetics 10, 134–140 (2009).

25. Bycroft, C. et al. The UK Biobank resource with deep phenotyping and genomic data. Nature 562, 203–209 (2018).

26. Weine, E., Smith, S.P., Knowlton, R.K. & Harpak, A. Tradeoffs in Modeling Context Dependency in Complex Trait Genetics. bioRxiv (2024).

27. Astore, C. & Gibson, G. Integrative polygenic analysis of the protective effects of fatty acid metabolism on disease as modified by obesity. Front Nutr 10, 1308622 (2023).

28. Borges, M.C. et al. Role of circulating polyunsaturated fatty acids on cardiovascular diseases risk: analysis using Mendelian randomization and fatty acid genetic association data from over 114,000 UK Biobank participants. BMC Med 20, 210 (2022).

29. Inouye, M. et al. Genomic Risk Prediction of Coronary Artery Disease in 480,000 Adults: Implications for Primary Prevention. J Am Coll Cardiol 72, 1883–1893 (2018).

30. Isgut, M., Sun, J., Quyyumi, A.A. & Gibson, G. Highly elevated polygenic risk scores are better predictors of myocardial infarction risk early in life than later. Genome Med 13, 13 (2021).

31. Fahed, A.C. & Natarajan, P. Clinical applications of polygenic risk score for coronary artery disease through the life course. Atherosclerosis 386, 117356 (2023).

32. Truong, B. et al. Modification of coronary artery disease clinical risk factors by coronary artery disease polygenic risk score. Med 5, 459–468 e3 (2024).

33. Ye, Y. et al. Interactions Between Enhanced Polygenic Risk Scores and Lifestyle for Cardiovascular Disease, Diabetes, and Lipid Levels. Circ Genom Precis Med 14, e003128 (2021).

34. Leinonen, J.T. et al. Genetic analyses implicate complex links between adult testosterone levels and health and disease. Commun Med (Lond) 3, 4 (2023).

35. Chen, H. et al. Association of serum lipids with inflammatory bowel disease: a systematic review and meta-analysis. Front Med (Lausanne) 10, 1198988 (2023).

36. Bai, L. et al. Computational drug repositioning of atorvastatin for ulcerative colitis. J Am Med Inform Assoc 28, 2325–2335 (2021).

37. Bigeh, A., Sanchez, A., Maestas, C. & Gulati, M. Inflammatory bowel disease and the risk for cardiovascular disease: Does all inflammation lead to heart disease? Trends Cardiovasc Med 30, 463–469 (2020).

38. Sleutjes, J.A.M. et al. Cardiovascular risk profiles in patients with inflammatory bowel disease differ from matched controls from the general population. Eur J Prev Cardiol 30, 1615–1622 (2023).

39. Hu, E.A. et al. Alcohol Consumption and Incident Kidney Disease: Results From the Atherosclerosis Risk in Communities Study. J Ren Nutr 30, 22–30 (2020).

40. Brittain, E.L. et al. Physical Activity and Incident Obesity Across the Spectrum of Genetic Risk for Obesity. JAMA Netw Open 7, e243821 (2024).

41. Thompson, M.D., Pirkle, C.M., Youkhana, F. & Wu, Y.Y. Gene-obesogenic environment interactions on body mass indices for older black and white men and women from the Health and Retirement Study. Int J Obes (Lond) 44, 1893–1905 (2020).

42. Tyrrell, J. et al. Gene-obesogenic environment interactions in the UK Biobank study. Int J Epidemiol 46, 559–575 (2017).

43. Nagpal, S., Gibson, G. & Marigorta, U.M. Pervasive Modulation of Obesity Risk by the Environment and Genomic Background. Genes (Basel) 9(2018).

44. Ding, Y. et al. Polygenic scoring accuracy varies across the genetic ancestry continuum. Nature 618, 774–781 (2023).

45. Hou, K., Xu, Z., Ding, Y., Harpak, A. & Pasaniuc, B. Calibrated prediction intervals for polygenic scores across diverse contexts. medRxiv (2023).

46. Urbut, S.M., Wang, G., Carbonetto, P. & Stephens, M. Flexible statistical methods for estimating and testing effects in genomic studies with multiple conditions. Nat Genet 51, 187–195 (2019).

47. Tucker-Drob, E.M., Briley, D.A. & Harden, K.P. Genetic and Environmental Influences on Cognition Across Development and Context. Curr Dir Psychol Sci 22, 349–355 (2013).

48. Khera, A.V. et al. Genome-wide polygenic scores for common diseases identify individuals with risk equivalent to monogenic mutations. Nature Genetics 50, 1219–1224 (2018).

49. Cook, R.J. & Sackett, D.L. The number needed to treat: a clinically useful measure of treatment effect. BMJ 310, 452–4 (1995).

50. Natarajan, P. et al. Polygenic Risk Score Identifies Subgroup With Higher Burden of Atherosclerosis and Greater Relative Benefit From Statin Therapy in the Primary Prevention Setting. Circulation 135, 2091–2101 (2017).

51. Williams, P. Retaining Race in Chronic Kidney Disease Diagnosis and Treatment. Cureus 15, e45054 (2023).

52. Hsu, J., Johansen, K.L., Hsu, C.Y., Kaysen, G.A. & Chertow, G.M. Higher serum creatinine concentrations in black patients with chronic kidney disease: beyond nutritional status and body composition. Clin J Am Soc Nephrol 3, 992–7 (2008).

53. Ku, E., McCulloch, C.E., Adey, D.B., Li, L. & Johansen, K.L. Racial Disparities in Eligibility for Preemptive Waitlisting for Kidney Transplantation and Modification of eGFR Thresholds to Equalize Waitlist Time. J Am Soc Nephrol 32, 677–685 (2021).

54. Westerman, K.E. & Sofer, T. Many roads to a gene-environment interaction. Am J Hum Genet 111, 626–635 (2024).

55. Kosiborod, M.N. et al. Semaglutide in Patients with Heart Failure with Preserved Ejection Fraction and Obesity. N Engl J Med 389, 1069–1084 (2023).

56. Perkovic, V. et al. Effects of Semaglutide on Chronic Kidney Disease in Patients with Type 2 Diabetes. N Engl J Med (2024).

57. Bick, A.G. et al. Genomic data in the All of Us Research Program. Nature 627, 340–346 (2024).

58. Purcell, S. et al. PLINK: a tool set for whole-genome association and population-based linkage analyses. Am J Hum Genet 81, 559–75 (2007).

59. Ge, T., Chen, C.-Y., Ni, Y., Feng, Y.-C.A. & Smoller, J.W. Polygenic prediction via Bayesian regression and continuous shrinkage priors. Nature Communications 10, 1776 (2019).

60. Loh, P.R. et al. Contrasting genetic architectures of schizophrenia and other complex diseases using fast variance-components analysis. Nat Genet 47, 1385–92 (2015).

